# Photoacoustic monitoring of angiogenesis predicts response to therapy in healing wounds

**DOI:** 10.1101/2021.10.13.21264867

**Authors:** Yash Mantri, Jason Tsujimoto, Brian Donovan, Christopher C. Fernandes, Pranav S. Garimella, William F. Penny, Caesar A. Anderson, Jesse V. Jokerst

## Abstract

Chronic wounds are a major health problem that cause the medical infrastructure billions of dollars every year. Chronic wounds are often difficult to heal and cause significant discomfort. Although wound specialists have numerous therapeutic modalities at their disposal, tools that could 3D-map wound bed physiology and guide therapy do not exist. Visual cues are the current standard but are limited to surface assessment; clinicians rely on experience to predict response to therapy. Photoacoustic (PA) ultrasound (US) is a non-invasive, hybrid imaging modality that can solve these major limitations. PA relies on the contrast generated by hemoglobin in blood which allows it to map local angiogenesis, tissue perfusion and oxygen saturation—all critical parameters for wound healing. This work evaluates the use of PA-US to monitor angiogenesis and stratify patients responding *vs*. not-responding to therapy. We imaged 19 patients with 22 wounds once a week for at least three weeks. Our findings suggest that PA imaging directly visualizes angiogenesis. Patients responding to therapy showed clear signs of angiogenesis and an increased rate of PA increase (p = 0.002). These responders had a significant and negative correlation between PA intensity and wound size. Hypertension was correlated to impaired angiogenesis in non-responsive patients. The rate of PA increase and hence the rate of angiogenesis was able to predict healing times within 30 days from the start of monitoring (power = 88%, alpha = 0.05) This early response detection system could help inform management and treatment strategies while improving outcomes and reducing costs.

## Introduction

Chronic wounds are a major health problem, but there are no tools to diagnose these wounds before they have erupted and/or evaluate deep tissue response to therapy.(1) Chronic wounds cost the United States medical infrastructure up to $100B/year with a single diabetic ulcer costing nearly $50,000 — these numbers will increase as the population ages.(2) To decrease costs and improve quality of life, the community needs tools to predict and monitor response to therapy. Unfortunately, gold standard methods are primarily based on visual inspection and cannot see beneath the skin surface — 3D mapping of physiology deep into the wound bed could better stratify wound risk and guide therapy but such tools do not exist. While the Braden/Norton scales and transcutaneous oximetry (TCOM) have shown promise, these systems offer an ensemble assessment of the affected area with no spatial details on the wound boundaries, wound depth, and interaction of wound with healthy tissue.(3, 4) Thus, the development of tools to map and measure imaging markers associated with wound risk and treatment response could have a major positive impact for patients with chronic wounds or at risk of developing such wounds.(5-7)

Ultrasound (US) imaging is non-invasive and rapid (8-13) and can make 3D maps of the wound. US is an affordable, high resolution, sensitive, non-ionizing, and real-time tool for imaging but its use is surprisingly rare in wound care despite being ideally suited to characterize soft tissue and bone surfaces.(14) Recently, we reported the use of US to assess wound size in 45 patients.(15) We also performed a longitudinal study of wound healing in patients who received allogenic skin grafts over a 110-day period. We showed that ultrasound imaging can predict wound exacerbation and tissue loss before it is seen by the eye.(15) However, ultrasound alone mostly provides *anatomic* information: There are few details on perfusion or oxygenation, which are critical to wound formation and wound healing. In contrast photoacoustic (PA) ultrasound is a “light in, sound out” technique versus conventional “sound in, sound out” ultrasound. Contrast in photoacoustic s is generated by differential absorption of light: hemoglobin and deoxyhemoglobin are common absorbers.(16-19) Thus, photoacoustic imaging can report tissue oxygenation and tissue perfusion.(20, 21) The same scan also collects standard ultrasound images.

Angiogenesis is the formation of new blood vessels from pre-existing vessels. It is well known that angiogenesis is crucial for wound healing.(22) The new blood vessels carry essential cytokines and oxygen for wound repair. Studies have shown that elevated glucose levels in diabetic patients hinders angiogenesis resulting in diabetic ulcer formation, poor wound healing, and limb loss.(23, 24) Treatment protocols such as hyperbaric oxygen therapy,(25) negative pressure wound therapy,(26) and debridement(27) can promote angiogenesis and improve healing outcomes. Hypertension can impair angiogenesis.(28) Hence, an early angiogenesis detection tool could help direct treatment protocols and drastically improve outcomes. Multi-photon microscopy techniques can visualize angiogenesis in vivo but these have micron-scale depth penetration. PA imaging is ideally suited for this application due to centimeter-scale depth penetration and the contrast generated by hemoglobin in blood vessels.(29, 30) Others have recently demonstrated the use of PA imaging to assess peripheral hemodynamic changes in humans.(31-35), and thus we were motivated to use photoacoustic imaging to visualize angiogenesis. This could help clinicians make early and better-informed decisions on whether a particular treatment regimen should be continued.

## Materials and Methods

### Patients

This study was performed in accordance with the ethical rules for human experimentation stated in the 1975 Declaration of Helsinki and approved by the University of California San Diego’s Human Research Protections Program (Institutional Review Board No. 191998 and 202019X). Informed written consent was acquired from all participants before scanning. Inclusion criteria were (i) age >18 years and be able to provide consent; (ii) wounds smaller than 15 cm^2^; (iii) patients must undergo a minimum of three scans spaced at least one week apart from each other. Exclusion criteria included (i) presence of secondary lesions at the wound site (e.g., melanomas); (ii) blood-borne diseases; (iii) orthopedic implants near the wound site. Nineteen patients (22 wounds) were recruited for this study at the UCSD Hyperbaric Medicine and Wound Care Center, Encinitas, CA, USA. **Table S1** describes the patient demographic.

All patients were scanned during a routine wound care visit. Patients were scanned once a week for at least 3 weeks. C.A.A. was the independent wound specialist and decided the treatment regimen for all patients blinded to the results of the scan. Before scanning, all wound dressings were removed per standard of care, and the wound area was cleaned using sterile saline. Surrounding healthy tissue was cleaned using alcohol swabs to prevent infection. A sterile CIV-Flex transducer cover (Product no. 921191, AliMed Inc., Dedham, MA, USA) was used for every scan to prevent cross contamination.

### Photoacoustic - Ultrasound Imaging

We used a commercially available LED-based photoacoustic imaging system (AcousticX from Cyberdyne Inc., Tsukuba, Japan). The AcousticX system uses two LED-arrays operating at 850 nm, pulse width 70 ns, and 4 kHz repetition rate. The 128-element linear ultrasound transducer operates at a central frequency of 7 MHz, bandwidth of 80.9%, and a 4 cm field of view. We used a custom hydrophobic gel pad from Cyberdyne Inc. and sterile ultrasound coupling gel (Aquasonic 100, Parker Laboratories Inc., Fairfield NJ, USA) for coupling with the wound surface. All images were acquired at 30 frames/s.

All wounds were scanned in a single sweep from inferior healthy tissue to wound region to superior healthy tissue. All scans were performed by hand, and thus frame alignment between scans was extremely difficult. Due to limitations in image exportation from the software, and to minimize misalignment effects between scans, we chose three representative frames from the central region of the wound for processing. Clinicians also report size and healing assessment from the wound’s center.(36) Furthermore, we matched the underlying bone pattern to compare similar spots over time. Y.M. acquired all the images.

### Image Processing

J.T. performed all the image processing and was blinded to wound photographs and healing times. J.T. only received US scans of the patients. All frames were reconstructed and visualized using the AcousticX software (Cyberdyne Inc.; Version 2.00.10). We exported 8-bit PA, B-mode, and overlayed coronal cross-section images. The images were further processed using Fiji, an ImageJ extension, version 2.1.0/1.53c. Data was plotted using Prism version 9.0.0. We drew custom regions-of-interest (ROIs) for every frame. We quantified changes in wound area, tissue regeneration, scar tissue development, and photoacoustic intensity as a function of time.

Wound area and tissue regeneration were quantified using a previously described method.(15) Briefly, we determined a dynamic baseline US intensity of healthy tissue for each patient. Areas with intensity lower or higher than baseline values were classified as wound and scar tissue respectively. Wound and scar area were measured using custom ROIs that fit the above classification criteria. Changes in PA intensity was measured using rectangular ROIs (4 cm wide x 1 cm deep). ROIs were drawn under the dermal layer (first 2 mm) hence avoiding PA signal from scabs and hyperpigmented regions of the skin. PA ROIs was made larger to cover the entire field of view of the transducer (4 cm). This is important so we did not miss any signs of angiogenesis from the periphery of the wound. ROIs for PA intensity measurements were also kept constant for all patients eliminating any concerns of inter-rater reliability. All US and PA quantification were carried out on the same frames.

### Statistics

We measured wound area and PA intensity in three frames for each scan. The error bars in each figure represent the standard deviation within these three frames. A simple linear regression was fit to the data measuring changes in imaging markers over time; 95% confidence intervals for these fits are shown in each figure. Furthermore, we plotted the rate of PA change per day *vs*. the healing time for the study population and fit a one-phase exponential decay curve to it. We used a Pearson correlation test to determine the correlation between the time to heal (days) versus rate of PA increase comparing the null hypothesis that there is no correlation versus there is a negative correlation between these two variables. The statistical analyses were conducted at alpha = 0.05. A power analysis was also performed on this data. An area under the curve – receiver operating characteristic (AUC – ROC) analysis was performed to study the classification of therapeutic responders vs non-responders.

## Results

Nineteen patients with 22 wounds were enrolled in this study. All patients underwent at least three scans spaced one week apart. We measured changes in wound area, PA intensity, and scar tissue formation over time. **Table S2** lists all the wound and relevant patient information. Nine wounds showed response to therapy. **Table 1** shows the clinical features of therapeutic responders and non-responders. Responders were patients who healed within 111 days.(37, 38) Hypertension was significantly (p = 0.0001) responsible for delayed healing. We noted no significant difference in other clinical features (age, sex, diabetes, smoking, body mass index (BMI), heart rate, blood pressure, and oxygen saturation) between the two groups. Extreme cases of wounds that had a swift, delayed and no response to therapy have been highlighted below.

**Table 1.** Clinical features of therapeutic responders (healing time < 111 days) and non-responders. This data is from 19 patients with 22 wounds. Values are mean ± SD or number of subjects (%). * Marks significant difference (p < 0.05).

| Category | Responders<br>(n = 9 wounds) | Non-Responders<br>(n = 13 wounds) | p<br>values |
| --- | --- | --- | --- |
| Age (years) | 68.1 ± 10.3 | 60.9 ± 15.1 | 0.31 |
| Sex (male) | 2 (29 %) | 9 (75 %) | 0.051 |
| Diabetes | 2 (22 %) | 5 (38 %) | 0.59 |
| Hypertension | 1 (11 %) | 11 (85 %) | 0.0001* |
| Smoker | 2 (29 %) | 4 (31 %) | 0.39 |
| BMI (kg/m<sup>2</sup>) | 28.2 ± 9.6 | 30.5 ± 5.9 | 0.63 |
| Heart rate (bpm) | 74 ± 12 | 92 ± 22 | 0.19 |
| Blood Pressure (systolic/diastolic)<br>(mm of Hg) | 147/80 ± 22/6 | 154/91 ± 30/22 | 0.78 |
| Oxygen saturation (%) | 98.4 ± 0.97 | 97.7 ± 1.4 | 0.31 |
| Rate of PA change<br>(Intensity (a.u.)/day) | 6698 ± 4217 | 2501 ± 2129 | 0.002* |

**Figure 1** shows wound healing and angiogenesis in a female in her 80’s (Subject ID: PN1) presenting with a chronic, left posterolateral ankle ulcer. PN1 healed in 66 days. Wound healing was visible *via* photographs within the first 29 days of treatment (**Figure 1 A-C**). US imaging showed a 33.3% reduction in wound size over 29 days from 0.48 cm^2^ – 0.32 cm^2^ (**Figure 1P**). The wound area reduced linearly as a function of time (R^2^ = 0.61). PA imaging showed the formation of new blood vessels on day 7 (**Figure 1H**). PA intensity increased linearly at a rate of 4217 ± 1336 intensity a.u./day as the wound healed (R^2^ = 0.50) (**Figure 1Q**). A sagittal maximum intensity projection (MIP) of the wound area showed angiogenesis into the wound bed (**Figure 1 M-O**). Unannotated version of Figure 1 can be found in the supporting information (**Figure S1**). **Figure 1R** shows a negative correlation between wound area and PA intensity (R^2^ = 0.95).

**Figure 1.**
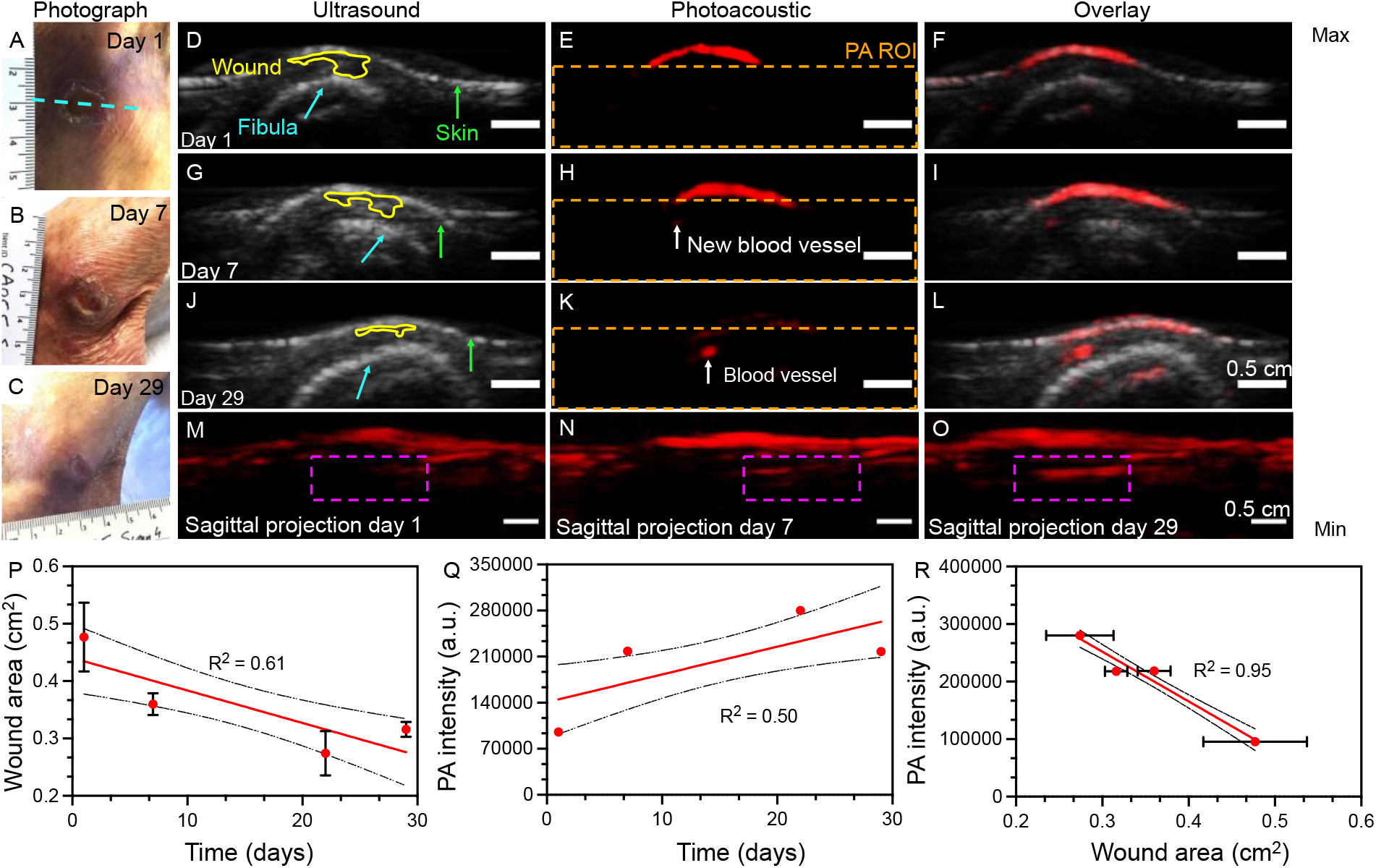
Photoacoustic imaging monitoring of angiogenesis in a healing wound. PN1, a female in her 80’s presenting with a chronic, left posterolateral ankle ulcer. **A-C**, Photographs showing the wound on days 1, 7, and 29 of the study. Blue dotted line indicates the imaging plane. **D-F, G-I**, and **J-L**, showing US, PA, and overlayed images of the wound on days 1, 7, and 29, respectively. Yellow ROI on the US outlines the wound. Green and blue arrows mark the skin surface and fibula respectively. White arrows (**H and K**) show new blood vessel formation *i*.*e*., angiogenesis. Orange outline marks the ROI for PA intensity measurement. **M-O**, show the sagittal maximum intensity projection of the wound on days 1, 7 and 29 showing new blood vessels invading the wound bed (Purple box). All scale bars are 0.5 cm. **P**, negative correlation between wound area and time suggests wound closure (R^2^ = 0.61). **Q**, Significant positive correlation between PA intensity and time suggests angiogenesis within the wound bed (R^2^ = 0.50). Rate of PA increase 4217 ± 1336 intensity a.u./day. **R**, PA intensity increases linearly as the wound heals suggesting that angiogenesis is correlated to wound closure (R^2^ = 0.95). Scale bars represent 0.5 cm. Error bars represent standard deviation in 3 representative frames from the center of the wound. Error bars for PA intensity in **Q** and **R** are too small to be shown.

**Figure 2** shows the progress of the wound healing indicators in PN2. PN2, was a male in his late 50’s presenting with a chronic left ankle ulcer following a severed Achilles tendon repair surgery. PN2 underwent three scans (day 1, 14, 28, 42) and took 292 days to heal. Photographs show tunneling of the wound under healthy surface tissue superior to the wound (**Figure 2D**). Blue dotted lines represent the imaging plane. It is important to note that tunneling wounds cannot be assessed non-invasively by the eye. **Figure 2A-C**, show wound progression over the 42-day study period. The wound tunnel showed 87% contraction by day 42 and wound area showed a strong negative correlation with treatment time (R^2^ = 0.89) (**Figure 2E**). More importantly, this patient showed the development of scar tissue by day 28 that was also mentioned in the doctor’s notes. Tissue was considered scarred if the mean US intensity was higher than healthy tissue baseline. Scar area was measured using custom ROIs with maximum size fitting the above criteria. Scar area and intensity increased linearly as a function of time (R^2^ = 0.43 and 0.61 respectively) (**Figure 2 F-G**). PA intensity in the wound area increased linearly at a rate of 4078 ± 534 intensity a.u./day, R^2^ = 0.85 (**Figure 2H**). PA intensity was negatively proportional to wound area (R^2^ = 0.64) (**Figure 2I**). Unannotated version of **Figure 2** can be found in the supporting information (**Figure S2**)

**Figure 2.**
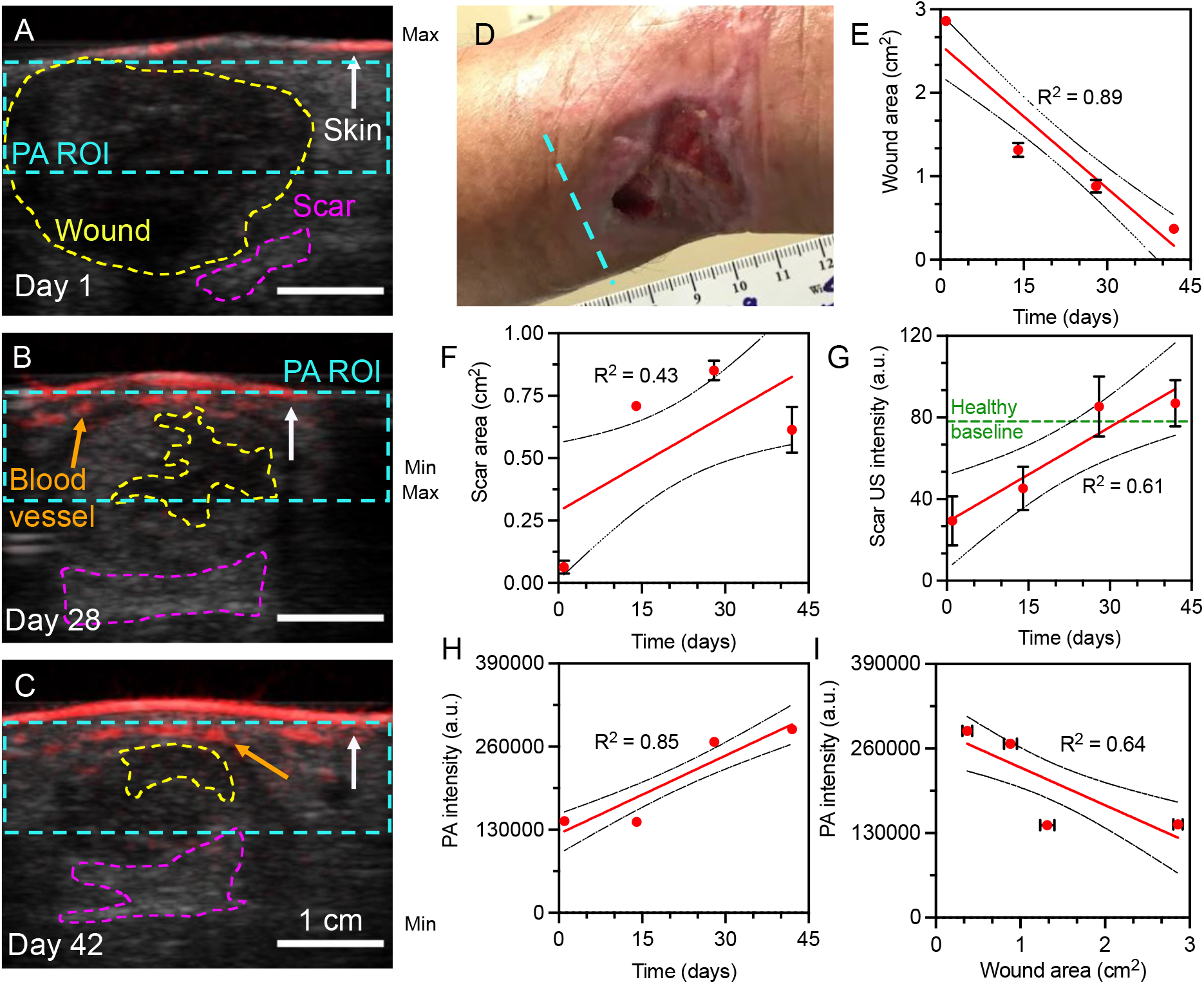
Tunneling wounds; wound closure, scar tissue development and angiogenesis. **A-C**, US-PA overlay of the wound on days 1, 28, and 42 of the study. Yellow, purple, and blue dotted lines in A-C represent wound, scar area, and PA ROI respectively. White and orange arrows represent skin surface and blood vessels respectively. **D**, Photographic image of wound in the left posterior ankle region. There is significant tunneling of the wound (not seen by eye). Blue dotted line in **D** indicates the relative imaging plane for panels A-C. **E**, 87% wound contraction is seen within 42-days. **F-G**, Scar tissue development is seen as hyperechoic regions at the wound bed. **H**, Significant increase in PA intensity over time indicates angiogenesis. **I**, A negative correlation between PA intensity and wound area suggests angiogenesis results in wound closure. Scale bars represent 1 cm. Error bars represent standard deviation in three frames at the center of the wound. Error bars for PA intensity in **H** and **I** are too small to be shown.

**Figure 3** shows progression in a non-healing wound. Subject PN3 was a female in her 70’s presenting with a stage III pressure ulcer on her left heel. PN3 took over 384 days to heal and was still receiving wound care during the preparation of this manuscript. PN3 underwent five scans over an 85-day period, and received standard wound care decided by the attending physician C.A.A. Photographs showed no visible contraction of the ulcer (**Figure 3 A, C, and E**). US imaging showed a 9.4% reduction in wound size over 85-days (**Figure 3G**). PA intensity increased by 4.2% during the same interval (**Figure 3H**). No clear signs of angiogenesis were visible at any point of the study. There was no significant correlation between PA intensity and wound area, R^2^ = 0.03 (**Figure 3I**). Unannotated version of **Figure 3** can be found in the supporting information (**Figure S3**). The data for individual patients showing changes in wound area, PA intensity as a function of time, and PA intensity *vs*. wound area are shown in the supporting information (**Figure S5-26**).

**Figure 3.**
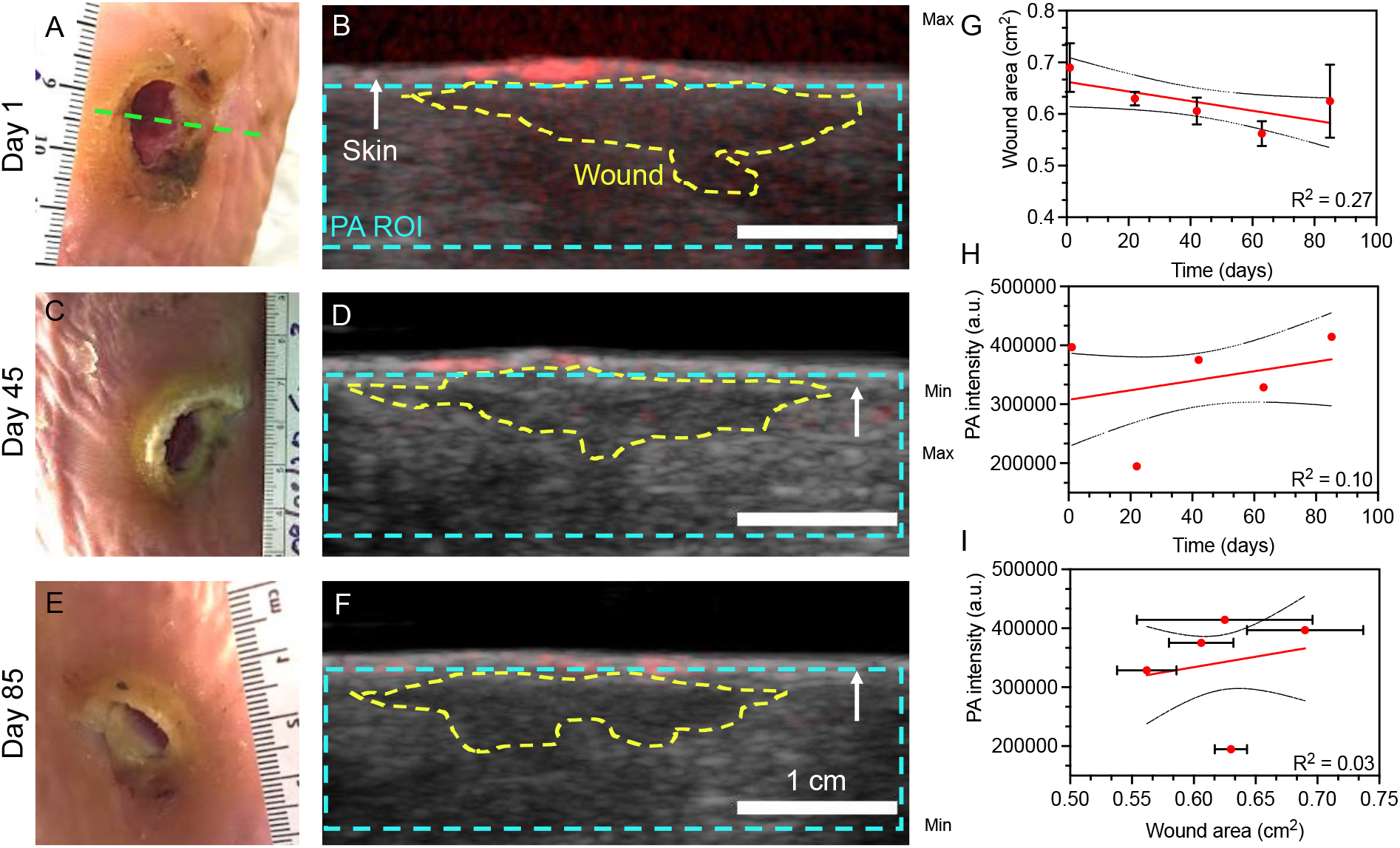
Wound progression in a non-responding patient. **A-B, C-D**, and **E-F**, photographs, and US – PA overlays on days 1, 45, and 85 of the study respectively. No significant changes in wound size can be seen in the pictures and US scans. Yellow and blue dotted line outlines the wound region and PA ROI used for processing. White arrow marks the skin surface. Green dotted line marks the imaging plane. **G**, mean wound area reduced by 9.4% in the 85-day period but this change was not statistically significant (R^2^ = 0.27). **H**, PA intensity in the wound increased at a rate of 807.7 ± 706.7 intensity a.u./day, R^2^ = 0.10 showing no significant correlation versus time. This suggests the absence of angiogenesis and the need for a different therapeutic approach. **I**, the plot of PA intensity *vs*. wound area showed no significant correlation (R^2^ = 0.03). Scale bars represent 1 cm. Error bars represent standard deviation in three frames at the center of the wound. Error bars for PA intensity in **H** and **I** are too small to be shown.

**Figure 4** shows population wide analysis for 17 patients with 20 wounds within the first 30 days of monitoring. The rate of PA increase was derived from the plot of PA intensity *vs*. time for each wound. Error bars represent the standard error of the slope. Healing times were noted from the patient charts as reported by the clinic and C.A.A. Two patients had scans more than 30 days apart and hence dropped from the analysis in **Figure 4A**. The full-length monitoring period for all patients can be found in the **Figure S4** that shows a similar trend as in **Figure 4A**. The minimum amount of time needed to classify a patient is 30 days. A one-phase exponential decay curve was fit to the data with an R^2^ = 0.76. The plateau was calculated to be 1738 intensity a.u./day. Wounds were classified into responders and non-responders using rate of PA change and healing time. 111-days was used as a cutoff for this classification based on previously reported values in literature.(37, 38) The green shaded region (n = 9 wounds) in **Figure 4A** shows wounds classified as responders to therapy. The other 11 wounds were classified as non-responders. A power analysis using the data in **Figure 4A** showed 80% power with an alpha of 0.05 with 17 wounds. With 20 wounds the power was 80% with p = 0.0009 (**Figure 4B**). Hence, the sample size was statistically sufficient to draw clinically significant conclusions. **Figure 4C** shows the AUC-ROC curves for discriminating responders *vs*. non-responders. Responders were patients with a rate of PA intensity increase greater than 1738 intensity a.u./day and healing time less than 111-days. The AUC-ROC value is 0.915, the highest among other reported wound prediction techniques (**Table 2**).

**Figure 4.**
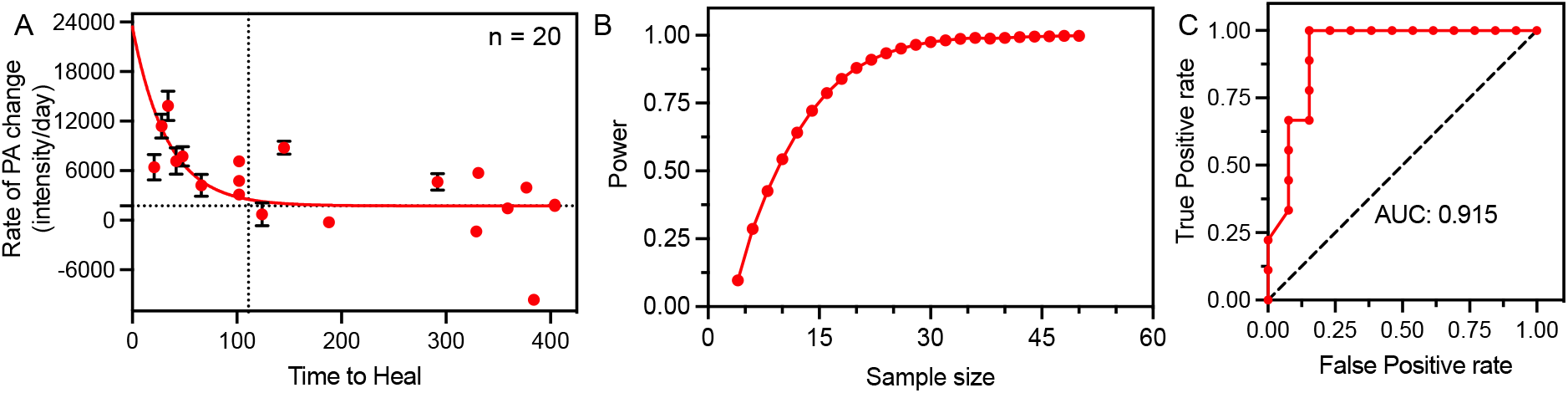
Photoacoustic imaging to predict wound healing and response to therapy. **A**, the rate of PA increase per day within the first 30 days is an effective imaging marker to predict wound healing time. Healing times reduce exponentially as a function of the rate of PA increase (n = 20). This could help classify patients as responders (green shaded area*) vs*. non-responders to a particular therapy. PA could help in the early identification of non-responders allowing clinicians to change their therapeutic approach and improve outcomes. **B**, Power analysis using the data used in panel A showed that 80% power with an alpha of 0.05 was achieved at n = 17. At n = 20 the power was 88%, p = 0.0009. Error bars in panel A represent standard error in the rate of PA change for each patient.

**Table 2.**
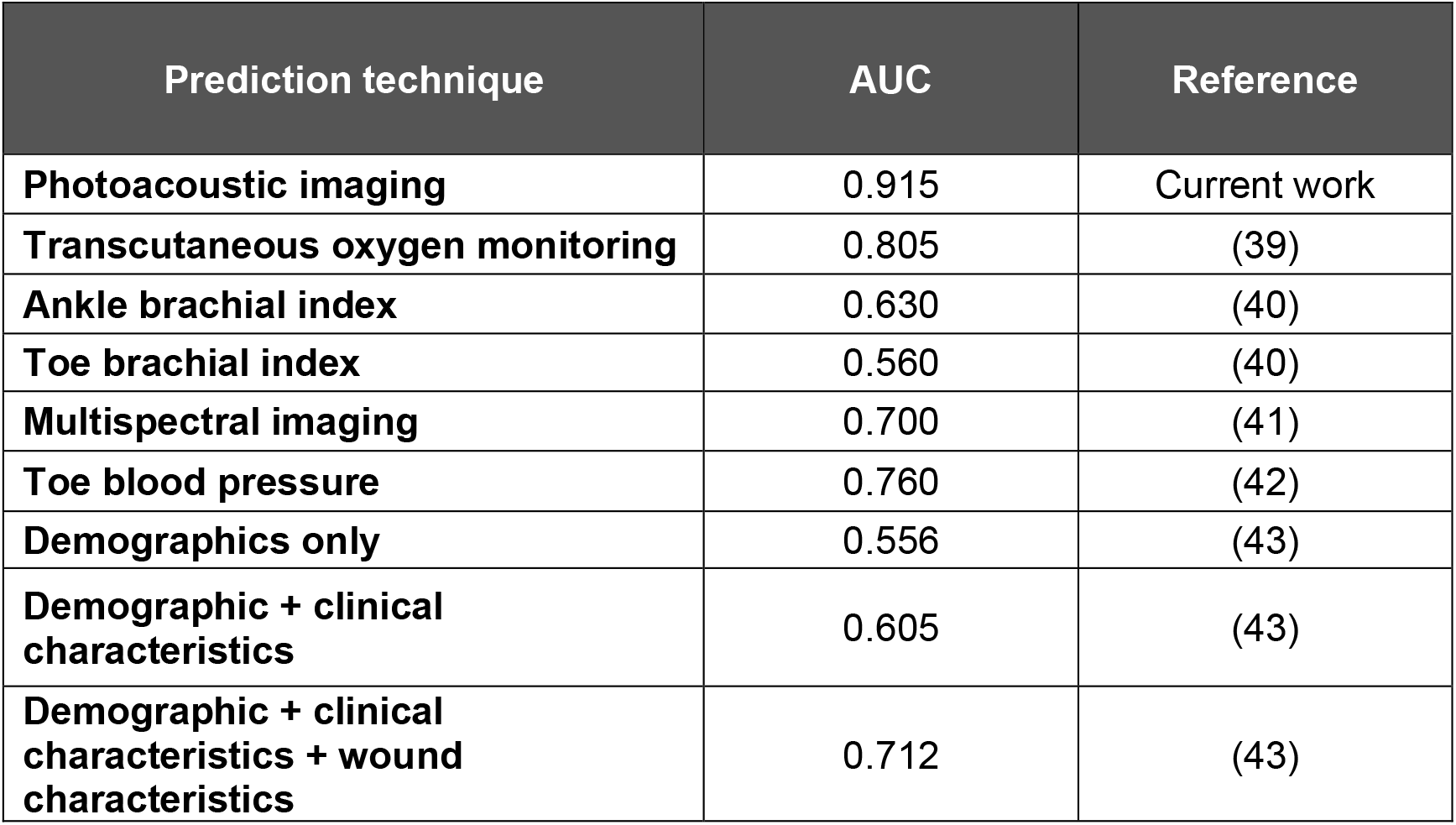
PA imaging has the highest AUC values compared to other commonly used wound healing prediction techniques.

| Prediction technique | AUC | Reference |
| --- | --- | --- |
| Photoacoustic imaging | 0.915 | Current work |
| Transcutaneous oxygen monitoring | 0.805 | (39) |
| Ankle brachial index | 0.630 | (40) |
| Toe brachial index | 0.560 | (40) |
| Multispectral imaging | 0.700 | (41) |
| Toe blood pressure | 0.760 | (42) |
| Demographics only | 0.556 | (43) |
| Demographic + clinical characteristics | 0.605 | (43) |
| Demographic + clinical characteristics + wound characteristics | 0.712 | (43) |

## Discussion

### Imaging Parameters

PA imaging is ideally suited to monitor local angiogenesis, perfusion, and oxygen saturation: These are all key parameters for wound healing.(44) Multiple studies have shown the use of PA tomography and microscopy to visualize the skin surface, superficial blood vessels, and blood flow with exceptional spatial resolution (< 100 µm, lateral resolution)(21, 45-48) The LED-based PA system used in this study has much lower spatial resolution and fluence but is also less expensive and more robust/portable compared to conventional high energy laser-based systems. It employs low-energy LED illumination operating under the maximum permissible exposure limit (2 – 9 µJ/cm^2^) with a lateral resolution between 550 – 590 µm.(49) Hand-held scans using the LED-based PA system allows easy mapping of wounds on contoured surfaces such as the ankle, making it ideal to visualize angiogenesis in complex wounds. The 850-nm excitation used in this study falls within the biological optical window and maximizes depth penetration while maintaining a relatively high signal-to-noise ratio (∼ 35 dB).(50) Limitations of this LED-based system include a small cache: The system acquires 500 – 1500 frames per scan but the processing software only exports 180 representative frames per scan (1 exported frame for every 8 acquired frames). Hence, there is a large loss of data unless one scans multiple small areas separately. The image exportation limited us to analyze only 3 representative frames from the center of the wound. Since the scans were performed using a hand-held transducer, it is difficult to know the precise distance between these representative frames. But analyzing the wound center minimizes the differences between scans.

One major strength of the study was that all image processing was carried out by J.T. who was blinded to the study and who had been trained in image classification by the attending physician. We used carefully considered criteria to define wound *vs*. scar *vs*. healthy tissue. Areas were classified as wound or scar tissue if the mean US intensity was lower or higher than healthy tissue baseline, respectively. Custom drawn ROIs were used to measure wound and scar area over time. Drawing custom ROIs can be extremely subjective (51) but we have shown good inter-rater reliability (mean bias 4.4%) in our previous work that used US to quantify tissue regeneration and wound closure in skin grafted patients.(15) The PA intensity was quantified using a rectangular ROI measure 4 cm wide and 1 cm deep and excluding the skin surface. We used the integrated density measurement which adds the intensity of all the pixels in the ROI instead of mean PA intensity. The use of integrated density reduces the effects of poor coupling, if any and provides an absolute value of PA intensity. The PA intensity ROI was maintained constant for all patients, eliminating concerns of subjectivity, and interferences due to skin tone.

### Clinical Significance

It is well established that angiogenesis is critical for wound healing. New blood vessels formed during the healing process deliver key cytokines and oxygen that reshape the wound matrix and result in wound closure.(44) Hence, angiogenesis can be a key imaging marker to predict response to therapy. The Centers for Medicare and Medicaid Services (CMS) in the United States re-evaluates coverage after 30 days from initial patient encounter. Patients needing advanced therapies need to be certified by the attending physician to enter a comprehensive plan of care in the medical record.(52) A recent high-powered study in 620,356 wounds showed that demographics, wound and clinical assessment could be used to predict wound healing in 84 days (AUC = 0.712, **Table 2**). But this is above the 30-day re-evaluation time limit set by CMS. (43)

The main clinical significance of this study is the ability to classify patients according to their response within 30 days from the start of therapy which aligns with the coverage re-evaluation time from CMS. Compared to other commonly employed techniques such as ankle brachial index, TCOM, *etc*., PA imaging is the best predictor for wound healing (AUC = 0.915, **Table 2**). PA classification could allow wound specialists to change their course of treatment if the wound is not responding to conventional treatment protocols.(15, 25, 53, 54) This would in turn improve outcomes, reduce amputations, healing time, and costs.

The rate of PA change is indicative of the rate of angiogenesis in the wound bed. The sagittal MIP (**Figure 1M-O**) confirms the formation of new blood vessels into the wound bed. Responding patients had a mean rate of PA change 6698 ± 4217 intensity (a.u.)/day that was significantly higher (p = 0.002) than non-responders (2501 ± 2129 intensity (a.u.)/day). Within the responding group, higher age and lower BMI were related to an increased rate of PA change (**Figure S27**). Age negatively impacts angiogenesis hence the age correlation is unexpected.(55, 56) The difference in treatment regimens like the use of cellularized tissue products to accelerate tissue regeneration could explain the age correlation. Blood pressure had no significant effect on the rate of PA change. Hypertension, diabetes, and smoking are also known to impair angiogenesis and hence wound repair.(28, 57, 58) The effect of hypertension on wound healing is visible in this cohort (**Table 1**): 12 of the 13 non-responsive wounds were hypertensive (92%), but only 1 of the 9 responsive wounds were hypertensive (14%). Hence non-hypertensive patients are more likely to develop new blood vessels and positively respond to therapy. Clinical factors alone can be used as a classifier but the use of PA imaging significantly improves prediction (**Table 2**).(43) A larger patient cohort could better illustrate the role of other risk factors that impair healing.

Traditionally, clinicians use surface cues such as color, temperature, odor, skin turgor, drainage, edema, and presence of devitalized tissue to assess wound health. In some cases, wound tunneling or cavitation can lengthen healing times and cause significant discomfort. Conventionally, probing tools are used to measure tunneling depth. Probing is invasive and often can lead to further tissue injury. Accurately and safely assessing tunneling wounds is therefore quite difficult visually. PN2 presents as an ideal example of a tunnelling wound to show the power of imaging over conventional wound assessment methods. The US was not only able to measure wound reduction (87% in 42 days), but also monitor scar tissue formation in the wound bed. Scar tissue presents as hyperechoic regions on the US due to its high fibrotic nature.(59) The addition of PA imaging allows us to visualize angiogenesis around the healing wound. Angiogenesis can be clearly seen in **Figure 2B-C** sandwiched in between the wound and skin surface. Deeper blood vessels can be seen on the US in **Figure 2C**, but these have very low PA signal due to reduced light penetration through tissue. The presence of a sterile sleeve between the transducer and skin surface also enhances light scattering, further reducing penetration depth. Using a higher wavelength of light could help visualize deeper vessels. The longer healing time compared to PN1 with similar rate of PA change can be attributed to the larger wound size, tunneling and a different treatment regimen compared to PN1. PN1s wound was limited to skin breakdown whereas PN2s wound had full thickness soft tissue involvement.

Secondary trauma, insufficient off-loading, poor wound dressing practices, and poor patient compliance can significantly impair wound healing and increase healing time.(60) Nevertheless, with 88% power in our study, we believe there is enough statistical significance to draw clinically relevant conclusions from this PA data. Future work in this field will look at employing oximetry-based PA measurements to measure local oxygen tension within the wound. It would also be interesting to study how PA imaging performs in conjunction with other prediction tools. The specialty of Hyperbaric Medicine could potentially benefit from this study. Such knowledge about oxygenation could potentially improve the use of hyperbaric oxygen treatment, indicating whether it should be initiated, continued, or halted. Patients not responding to therapy can then be more efficiently directed to other wound treatment interventions or therapeutic modalities. Furthermore motion-compensation and deep learning algorithms could improve image stability, quality, and streamline image processing.(61, 62)

## Conclusions

Angiogenesis is a key imaging marker for wound healing. PA-US imaging can be used to measure wound size, rate of angiogenesis, and scar tissue formation. A study of 19 patients with 22 wounds revealed that there is an inverse correlation between wound area and PA intensity. An increase in PA intensity correlates with wound closure due to the formation of new blood vessels. 3D MIP images confirmed blood vessel infiltration into the wound bed. Non-healing wounds showed no correlation between PA intensity and wound area. A higher rate of PA increase was associated with an exponential reduction in healing times. Finally, PA imaging could be used to classify therapy responders and non-responders within 30-days from the start of treatment. With an AUC value of 0.915, PA imaging is the best wound prediction technique. This work could have clinical significance in helping doctors make more informed and early decisions about whether treatment should be initiated, continued, altered, or halted. And suggest the need for more comprehensive Medicare coverage for non-responsive patients hence improving outcomes and reducing costs.

## Data Availability

All data produced in the present study are available upon reasonable request to the authors

## Acknowledgments

We acknowledge support from the National Institutes of Health through Grant R21 AG065776 as well as internal funds from the University of California San Diego, under the Galvanizing Engineering in Medicine Program. Y.M. acknowledges help from Brandon Brodish, RN, Luz Amezquita, RN, Starr Nimeth, LVN, and the entire team at the Hyperbaric Medicine and Wound Healing Center, University of California San Diego, Encinitas, CA, USA.

## List of Abbreviations

AUC-ROC: Area Under the Curve – Receiver Operating Characteristic
BMI: Body Mass Index
CMS: Centers for Medicare & Medicaid Services
MIP: Maximum Intensity Projection
PA: Photoacoustic
ROI: Region of Interest
TCOM: Transcutaneous Oximetry
US: Ultrasound

## Supporting Information

The supporting information contains patient wound descriptions and wound area *vs*. time, PA intensity *vs*. time, and PA intensity *vs*. wound area for all 22 wounds. Unannotated versions of Figure 1, 3, and 4.

