## Supplementary material for "Photoacoustic monitoring of angiogenesis predicts response to therapy in healing wounds": Table S1

**Optoacoustic imaging, Angiogenesis, Neovascularization, Healing time, Wound healing, Wound prediction, Early diagnosis**

**Abstract**

Chronic wounds are a major health problem that cause the medical infrastructure billions of dollars every year. Chronic wounds are often difficult to heal and cause significant discomfort. Although wound specialists have numerous therapeutic modalities at their disposal, tools that could 3D-map wound bed physiology and guide therapy do not exist. Visual cues are the current standard but are limited to surface assessment; clinicians rely on experience to predict response to therapy. Photoacoustic (PA) ultrasound (US) is a non-invasive, hybrid imaging modality that can solve these major limitations. PA relies on the contrast generated by hemoglobin in blood which allows it to map local angiogenesis, tissue perfusion and oxygen saturation—all critical parameters for wound healing. This work evaluates the use of PA-US to monitor angiogenesis and stratify patients responding *vs.* not-responding to therapy. We imaged 19 patients with 22 wounds once a week for at least three weeks. Our findings suggest that PA imaging directly visualizes angiogenesis. Patients responding to therapy showed clear signs of angiogenesis and an increased rate of PA increase (p = 0.002). These responders had a significant and negative correlation between PA intensity and wound size. Hypertension was correlated to impaired angiogenesis in non-responsive patients. The rate of PA increase and hence the rate of angiogenesis was able to predict healing times within 30 days from the start of monitoring (power = 88%, alpha = 0.05) This early response detection system could help inform management and treatment strategies while improving outcomes and reducing costs.

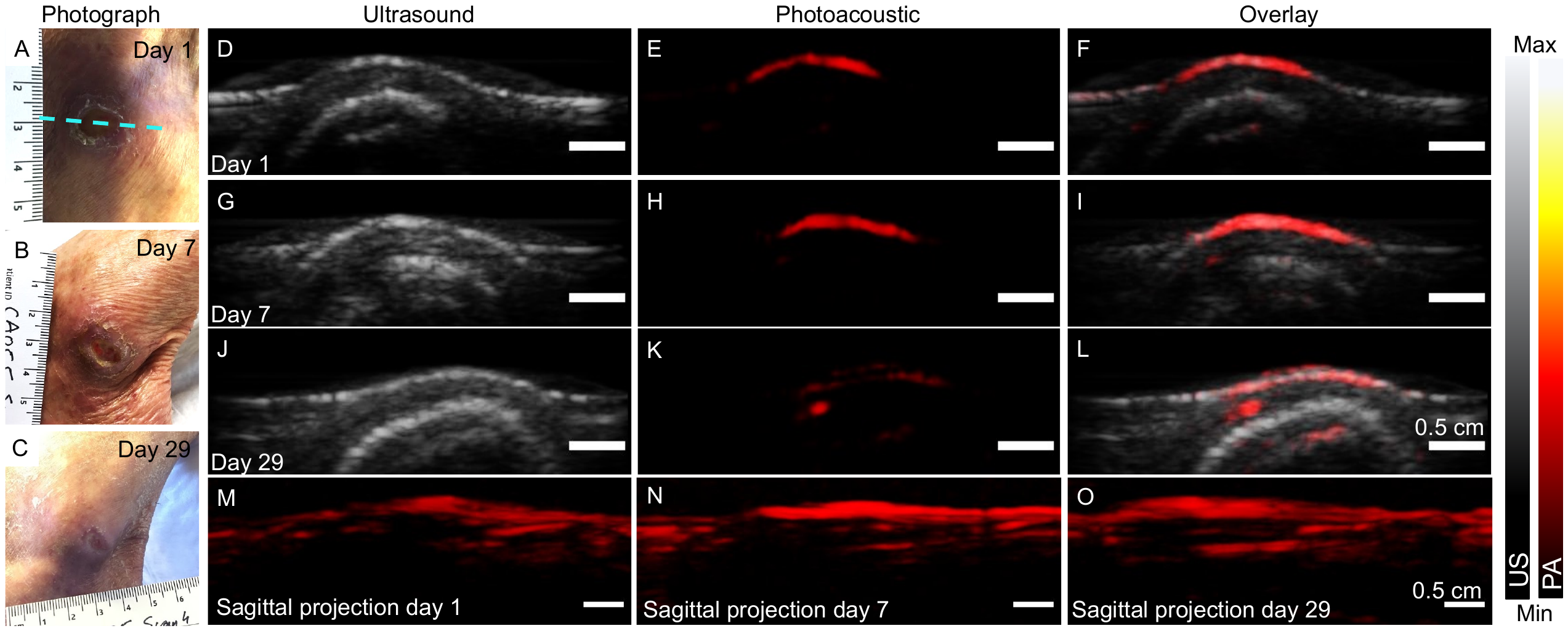

**Figure S1.** Unannotated version of Figure 1. Blue dotted line marks the imaging plane.

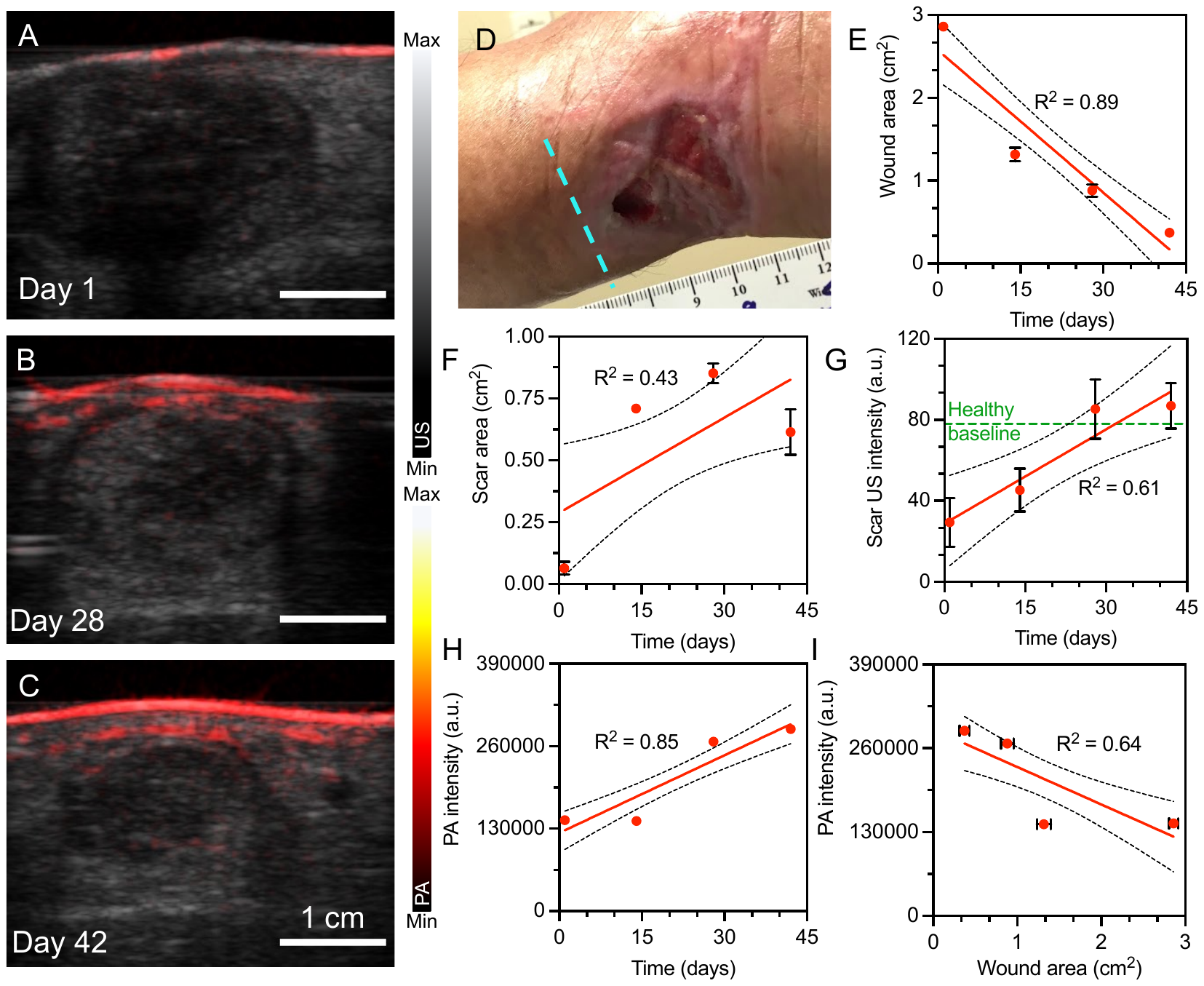

**Figure S2.** Unannotated version of Figure 3. Blue dotted line marks the imaging plane.

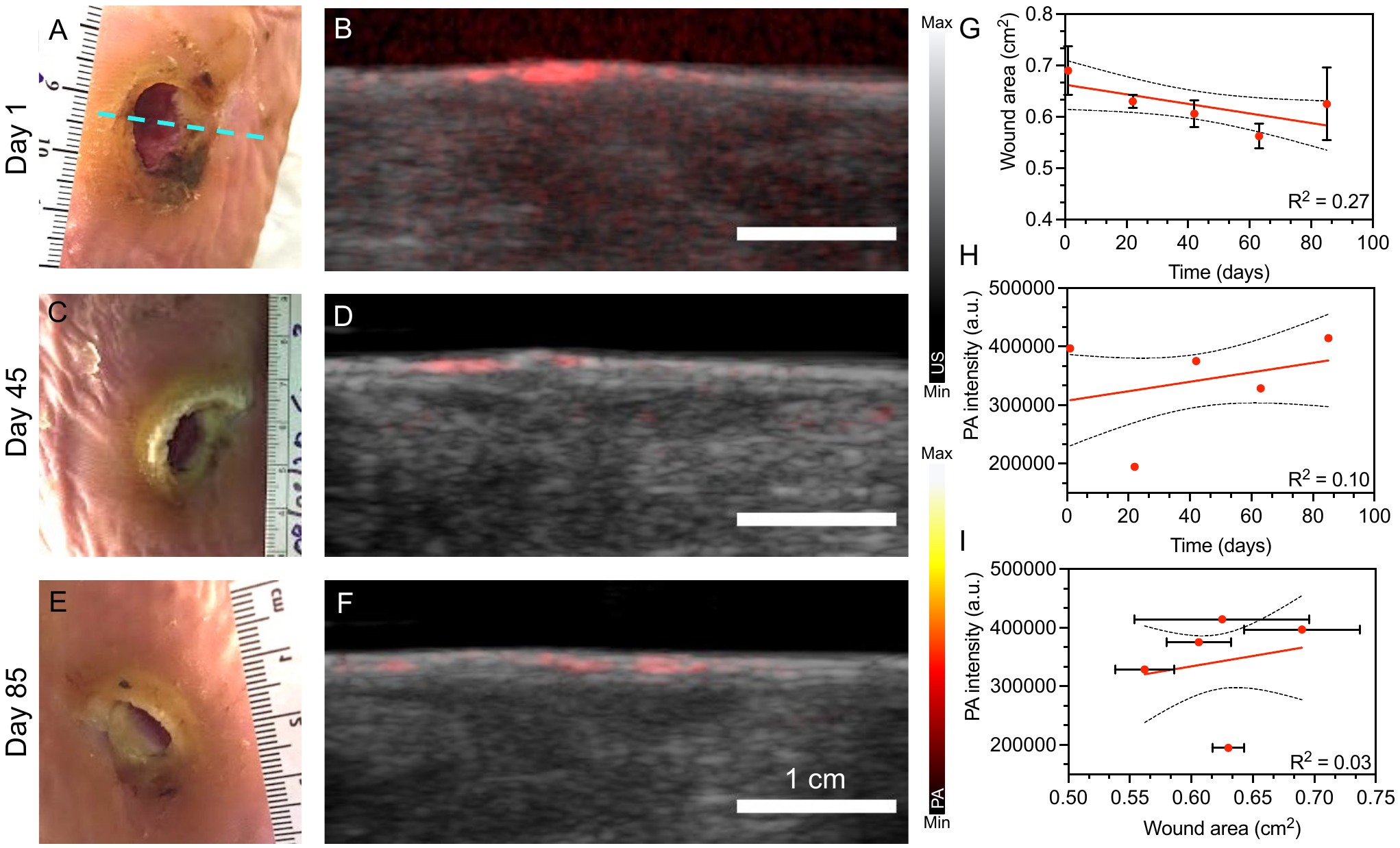

**Figure S3.** Unannotated version of Figure 4. Blue dotted line marks the imaging plane.

| **Category** | **Distribution** |
| --- | --- |
| **Total participants** | 19 |
| **Number of scanning events range** | 3 – 11 scans |
| **Monitoring time range for individual patients** | 21 – 112 days |
| **Average age (years)** | 63.5 ± 14.4 |
| **Sex (Male / Female)** | 11 / 8 |
| **Body mass index (kg/m^2^)** | 29.4 ± 7.4 |
| **Diabetes (Y / N)** | 8 / 11 |
| **Hypertension (Y / N)** | 12 / 7 |
| **Smoker (Y/N)** | 8 / 11 |

**Table S1.** Patient demographic distribution.

| **Patient Number** | **Age**  **Range**  **(yrs)** | **Sex**  **(M/F)** | **Number of Scans**  **(Study period in days)** | **Wound Description** | **Healing Time (Days)** | **Hypertensive**  **(Y/N)** | **Response to therapy (Y/N)** |
| --- | --- | --- | --- | --- | --- | --- | --- |
| **PN1** | 81-85 | F | 4 (29) | Left posterolateral ankle | 66 | N | Y |
| **PN2** | 56-60 | M | 4 (42) | Left posterior ankle | 292 | Y | N |
| **PN3** | 66-70 | F | 5 (85) | Left heel | 384 | Y | N |
| **PN4** | 66-70 | M | 6 (112) | Right Achilles ulcer | 411* | Y | N |
| **PN5** | 26-30 | M | 3 (118) | Inferior to the left knee | 357 | Y | N |
| **PN6** | 76-80 | M | 11 (110) | Inferior to the left knee | 124 | Y | N |
| **PN7 – WA** | 71-75 | M | 3 (21) | Left dorsal ankle | 404* | Y | N |
| **PN7 – WB** | 71-75 | M | 3 (21) | Left dorsal heel | 404* | Y | N |
| **PN8** | 56-60 | M | 4 (51) | Greater toe of right foot | 331 | Y | N |
| **PN9 – WA** | 56-60 | F | 4 (35) | Lower anterior left leg | 102 | N | Y |
| **PN9 – WB** | 56-60 | F | 4 (35) | Lower medial left leg | 102 | N | Y |
| **PN9 – WC** | 56-60 | F | 4 (35) | Lower left leg | 102 | N | Y |
| **PN10** | 71-75 | M | 7 (99) | Left posterior leg | 329 | Y | N |
| **PN11** | 76-80 | F | 6 (98) | Lateral left ankle | 188 | Y | N |
| **PN12** | 31-35 | M | 10 (106) | Lower anterior left leg | 145 | Y | N |
| **PN13** | 66-70 | F | 4 (34) | Toe of left foot | 34 | N | Y |
| **PN14** | 61-65 | M | 4 (77) | Left plantar great toe | 377* | Y | N |
| **PN15** | 51-55 | M | 4 (76) | Mid-lateral right foot | 359 | N | N |
| **PN16** | 61-65 | F | 3 (21) | Right lower leg | 21 | N | Y |
| **PN17** | 81-85 | F | 4 (28) | Right lower leg | 28 | N | Y |
| **PN18** | 56-60 | M | 6 (42) | Right medial ankle | 42 | Y | Y |
| **PN19** | 66-70 | M | 3 (22) | Right plantar region | 48 | N | Y |

**Table S2.** Patient and wound information. Green highlights indicate patients who are non-hypertensive and show good response to therapy. * Indicates patients who were still receiving wound care at the time of submission. WA, B, and C, denote different wounds on the same patient.

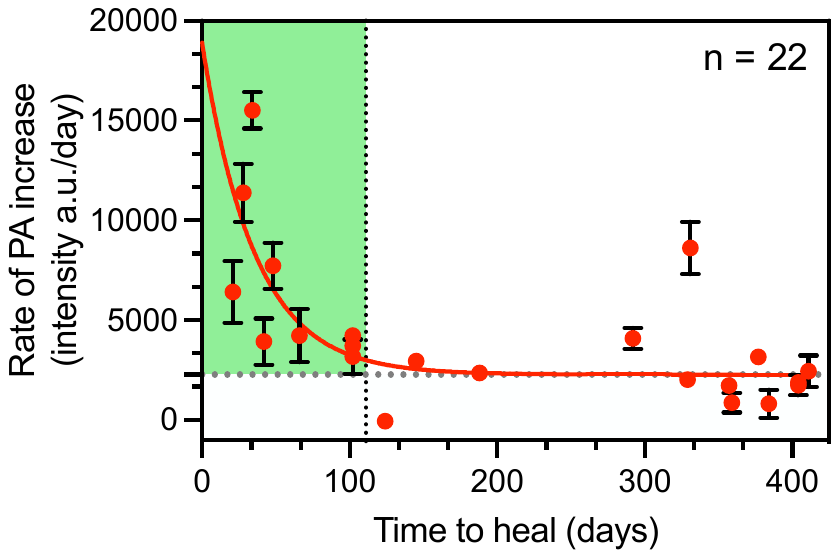

**Figure S4.** The rate of PA increase per day for the full study period is also an effective imaging biomarker to predict wound healing time. Healing times reduce exponentially as a function of the rate of PA increase (n = 22, R^2^ = 0.49)

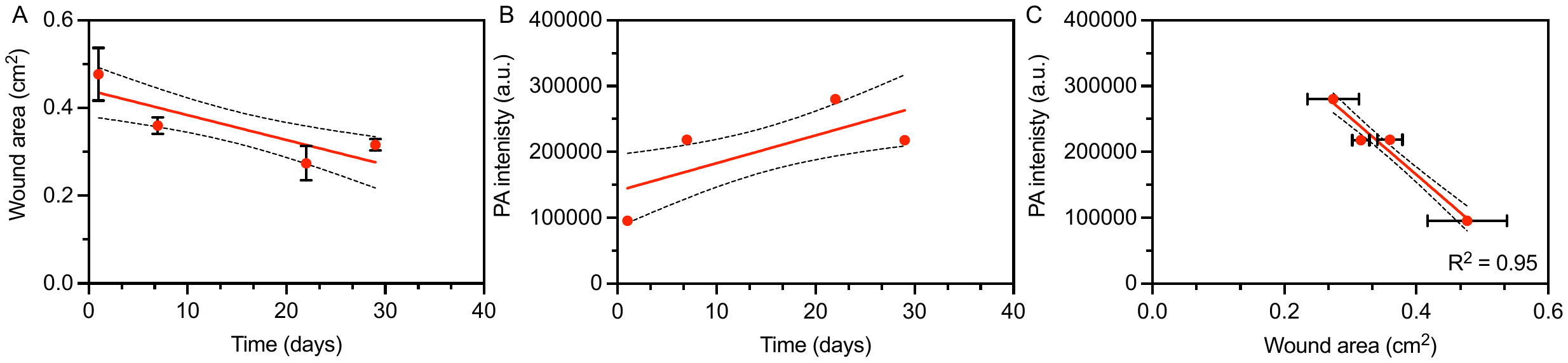

**Figure S5. A,** Wound area *vs.* time, **B**, PA intensity *vs.* time, and **C**, PA intensity *vs.* Wound area for PN1.

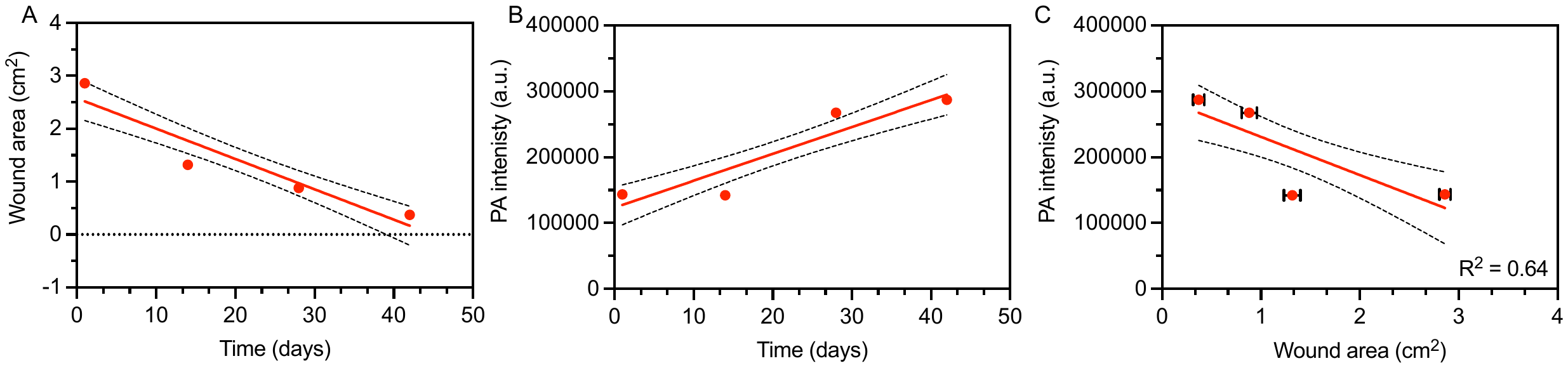

**Figure S6. A,** Wound area *vs.* time, **B**, PA intensity *vs.* time, and **C**, PA intensity *vs.* Wound area for PN2.

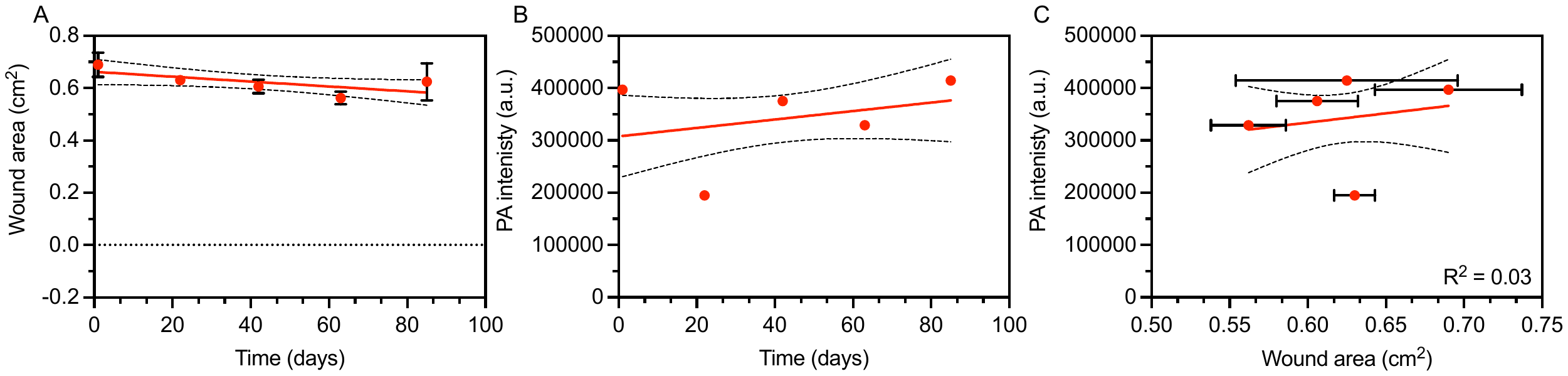

**Figure S7. A,** Wound area *vs.* time, **B**, PA intensity *vs.* time, and **C**, PA intensity *vs.* Wound area for PN3.

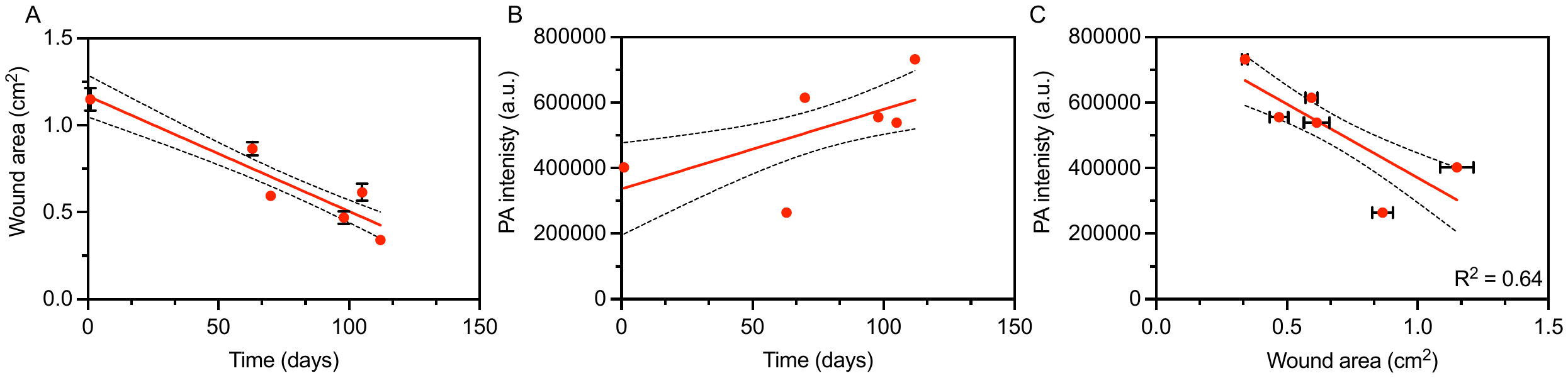

**Figure S8. A,** Wound area *vs.* time, **B**, PA intensity *vs.* time, and **C**, PA intensity *vs.* Wound area for PN4.

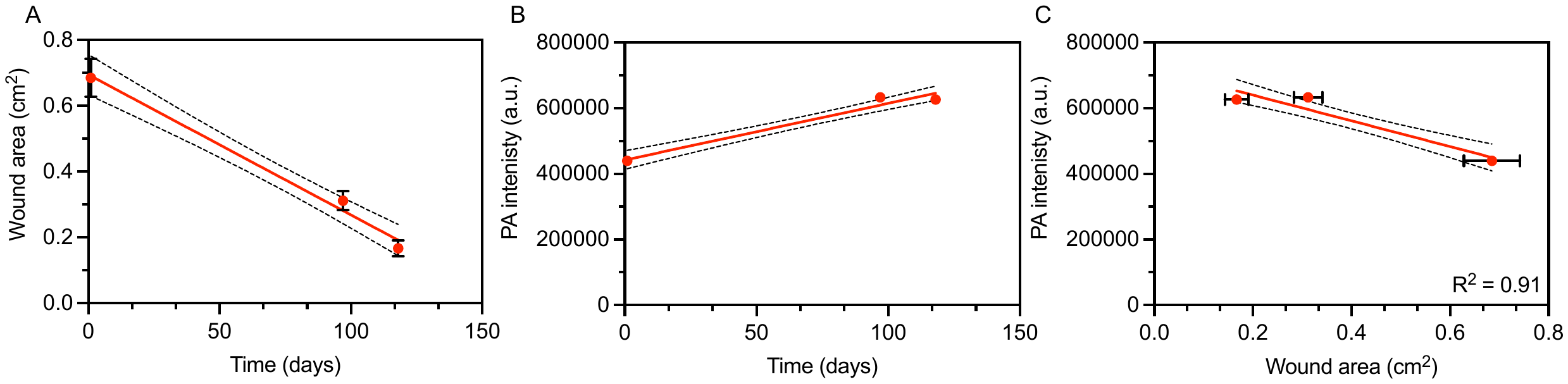

**Figure S9. A,** Wound area *vs.* time, **B**, PA intensity *vs.* time, and **C**, PA intensity *vs.* Wound area for PN5.

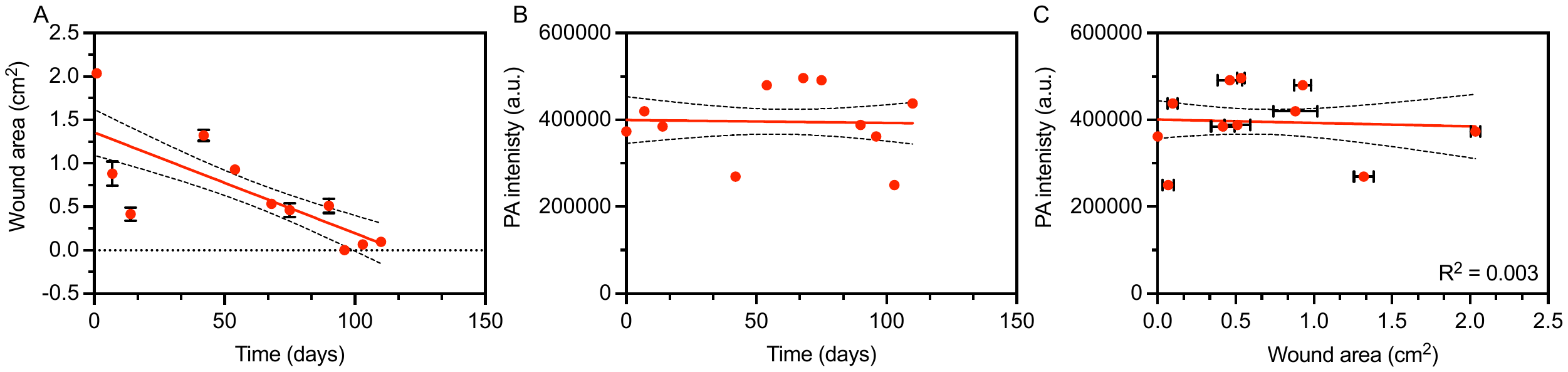

**Figure S10. A,** Wound area *vs.* time, **B**, PA intensity *vs.* time, and **C**, PA intensity *vs.* Wound area for PN6.

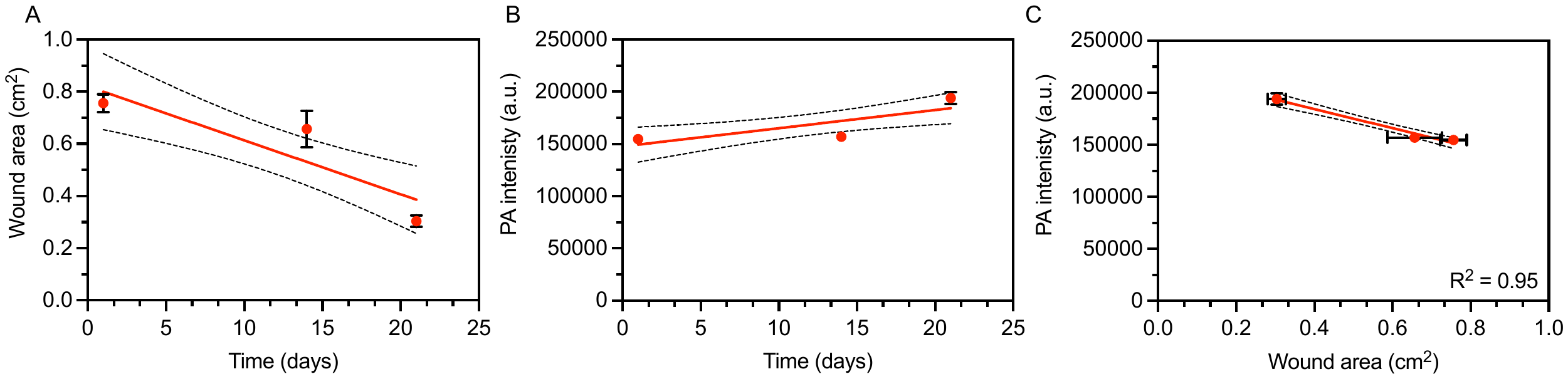

**Figure S11. A,** Wound area *vs.* time, **B**, PA intensity *vs.* time, and **C**, PA intensity *vs.* Wound area for PN7 – WA.

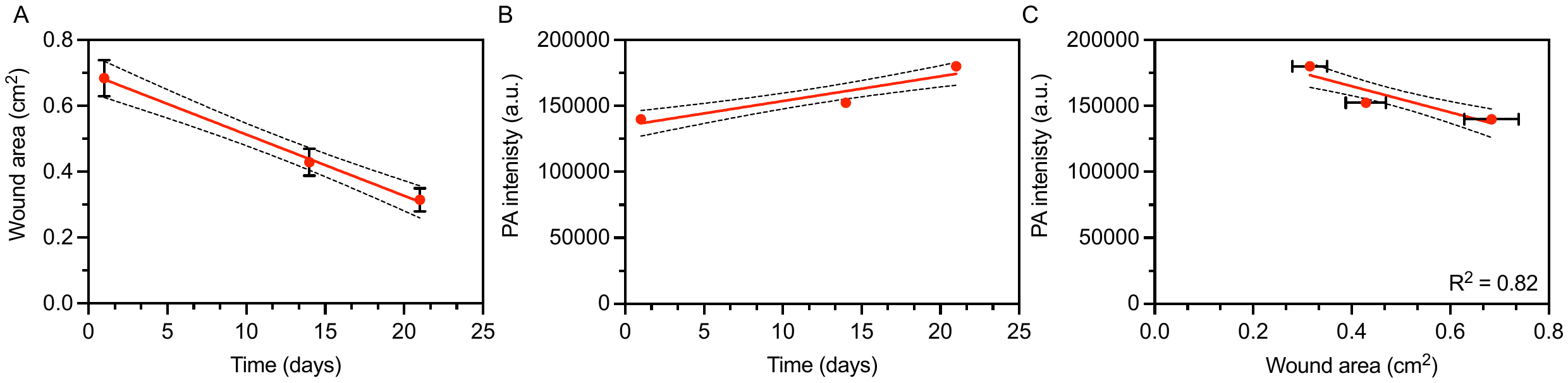

**Figure S12. A,** Wound area *vs.* time, **B**, PA intensity *vs.* time, and **C**, PA intensity *vs.* Wound area for PN7 – WB.

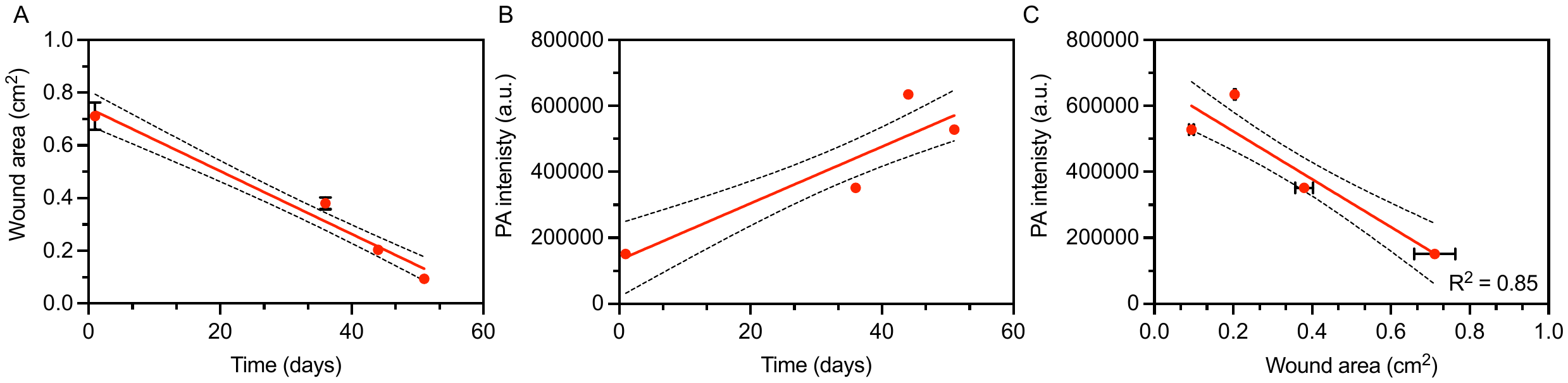

**Figure S13. A,** Wound area *vs.* time, **B**, PA intensity *vs.* time, and **C**, PA intensity *vs.* Wound area for PN8.

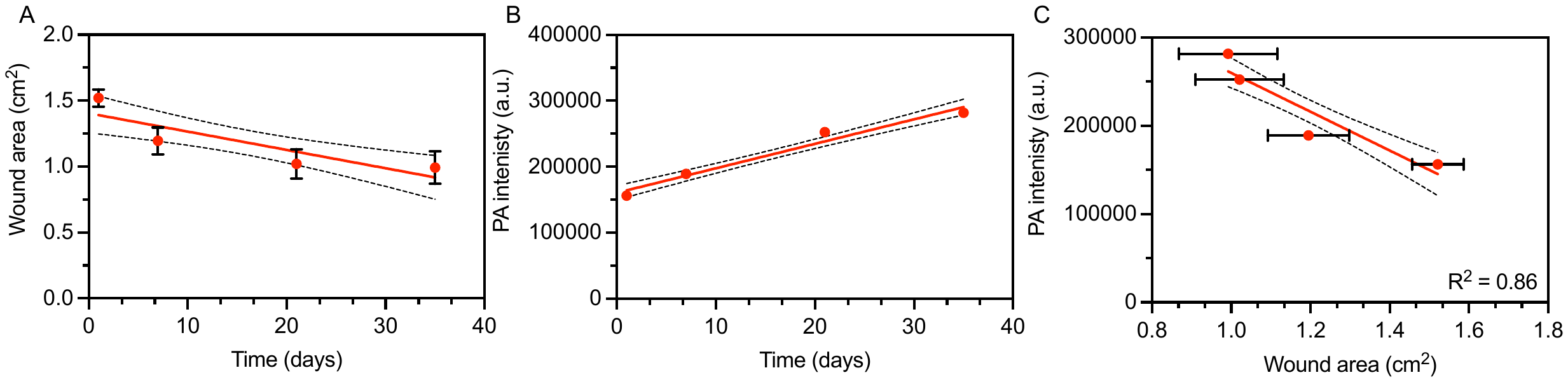

**Figure S14. A,** Wound area *vs.* time, **B**, PA intensity *vs.* time, and **C**, PA intensity *vs.* Wound area for PN9 – WA.

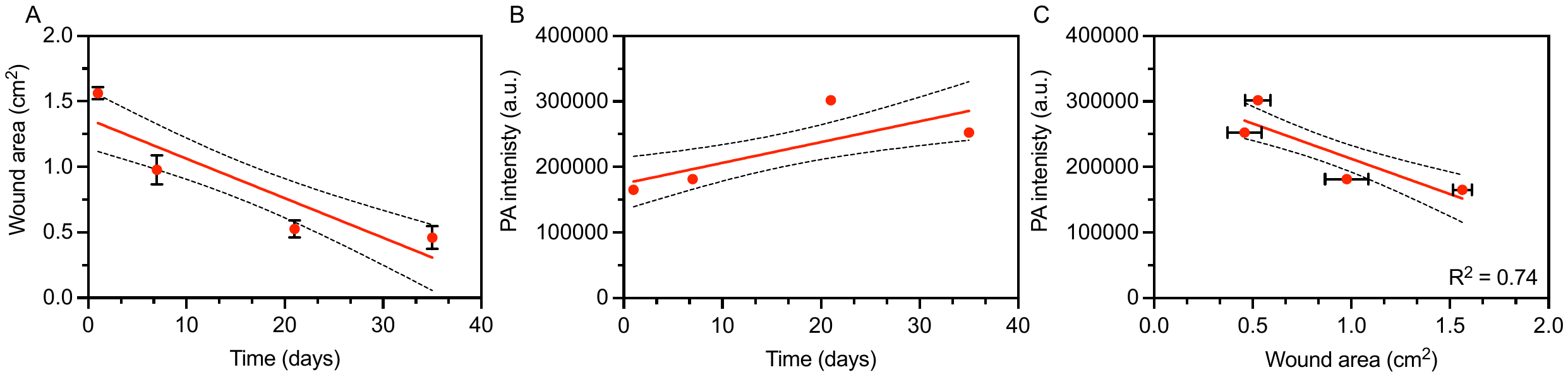

**Figure S15. A,** Wound area *vs.* time, **B**, PA intensity *vs.* time, and **C**, PA intensity *vs.* Wound area for PN9 – WB.

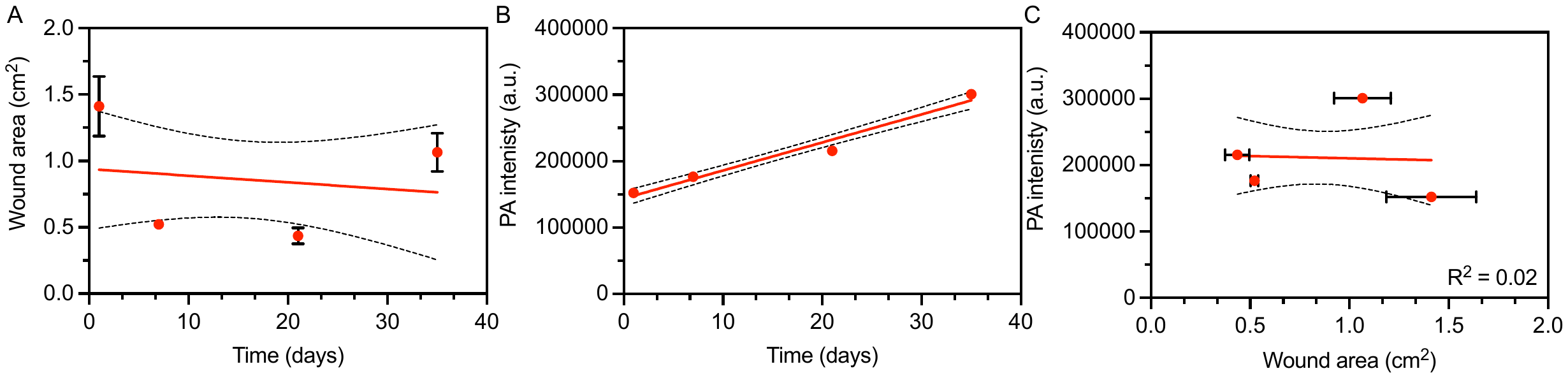

**Figure S16. A,** Wound area *vs.* time, **B**, PA intensity *vs.* time, and **C**, PA intensity *vs.* Wound area for PN9 – WC.

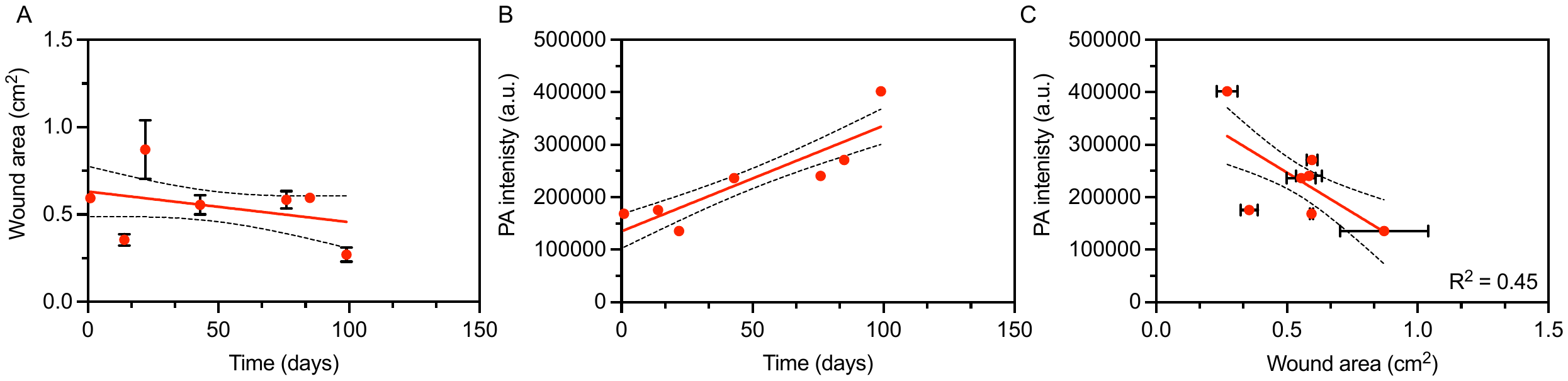

**Figure S17. A,** Wound area *vs.* time, **B**, PA intensity *vs.* time, and **C**, PA intensity *vs.* Wound area for PN10.

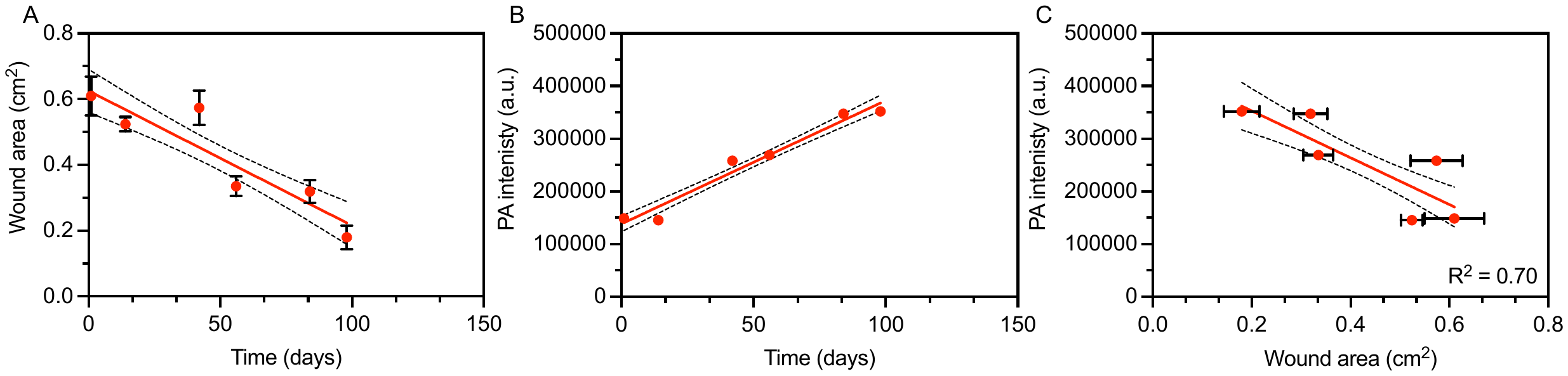

**Figure S18. A,** Wound area *vs.* time, **B**, PA intensity *vs.* time, and **C**, PA intensity *vs.* Wound area for PN11.

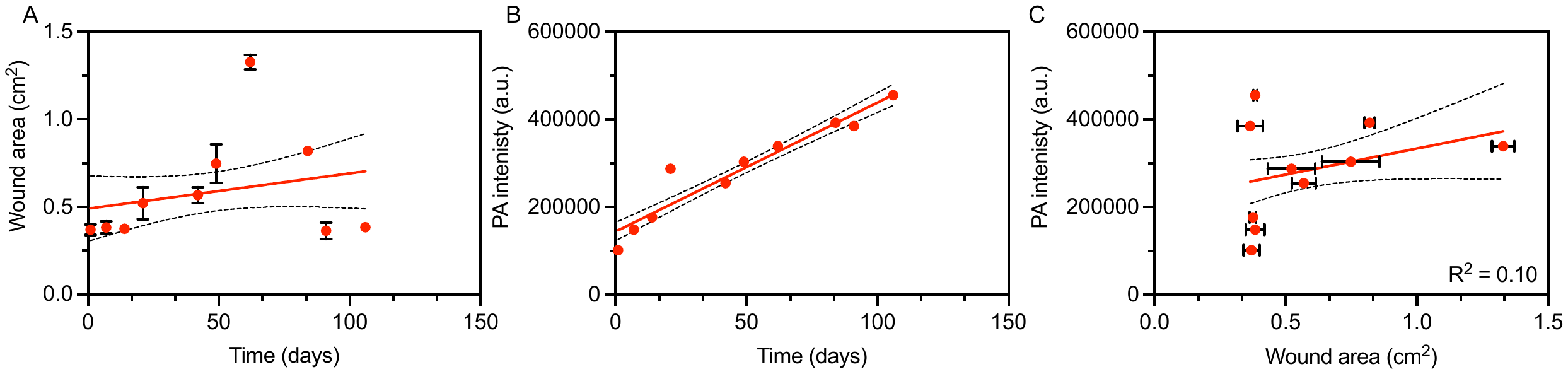

**Figure S19. A,** Wound area *vs.* time, **B**, PA intensity *vs.* time, and **C**, PA intensity *vs.* Wound area for PN12.

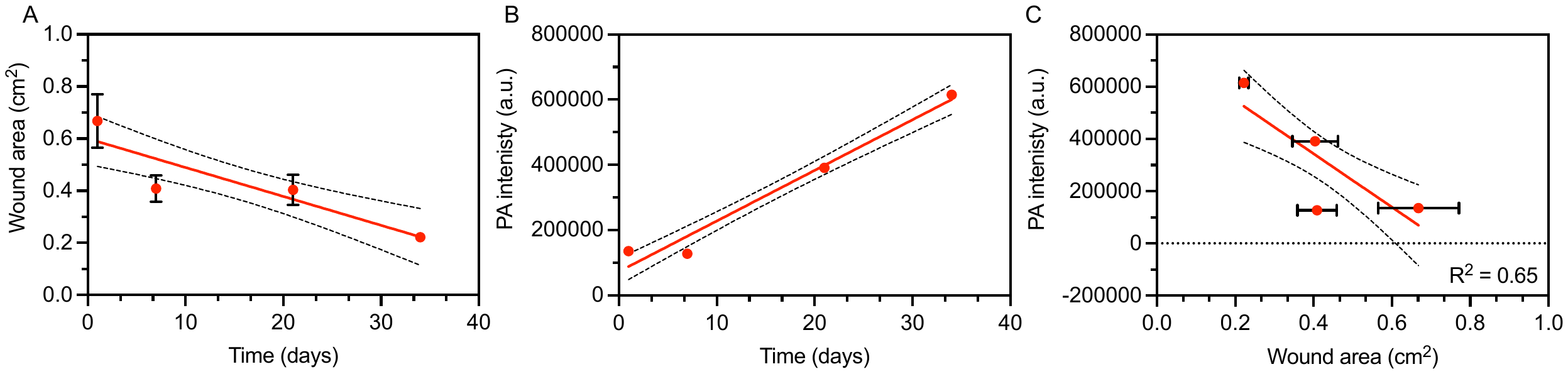

**Figure S20. A,** Wound area *vs.* time, **B**, PA intensity *vs.* time, and **C**, PA intensity *vs.* Wound area for PN13.

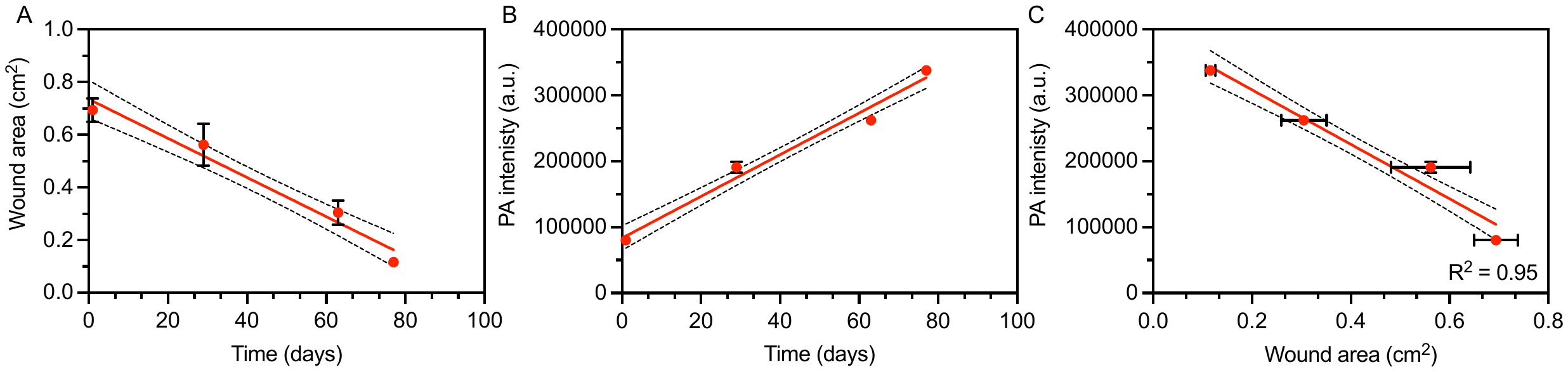

**Figure S21. A,** Wound area *vs.* time, **B**, PA intensity *vs.* time, and **C**, PA intensity *vs.* Wound area for PN14.

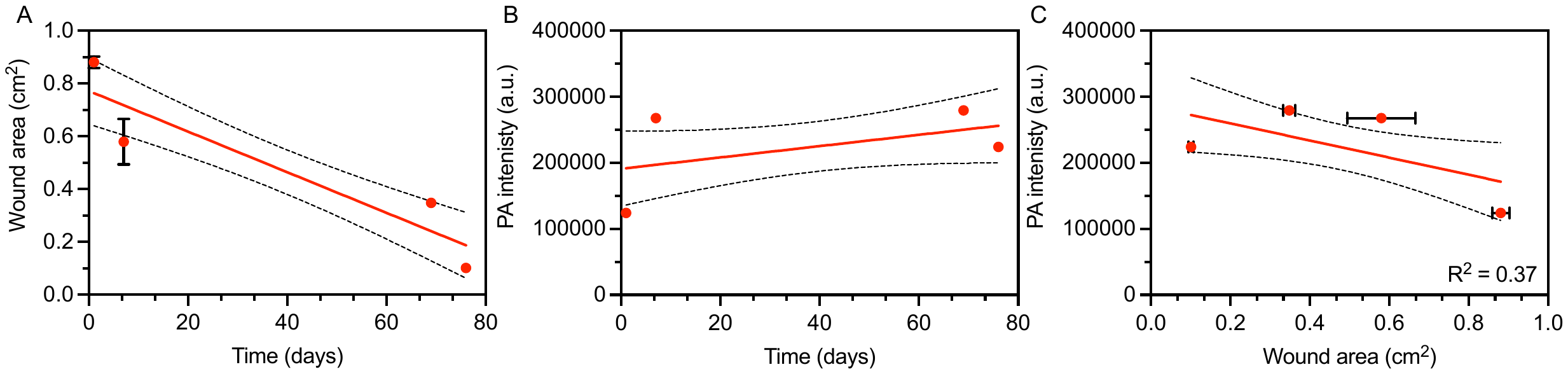

**Figure S22. A,** Wound area *vs.* time, **B**, PA intensity *vs.* time, and **C**, PA intensity *vs.* Wound area for PN15.

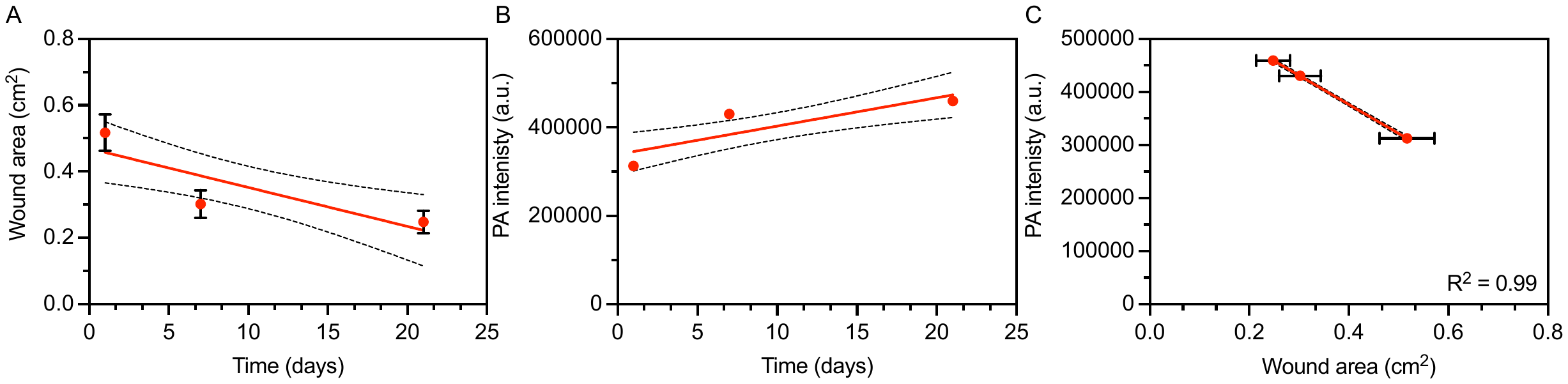

**Figure S23. A,** Wound area *vs.* time, **B**, PA intensity *vs.* time, and **C**, PA intensity *vs.* Wound area for PN16.

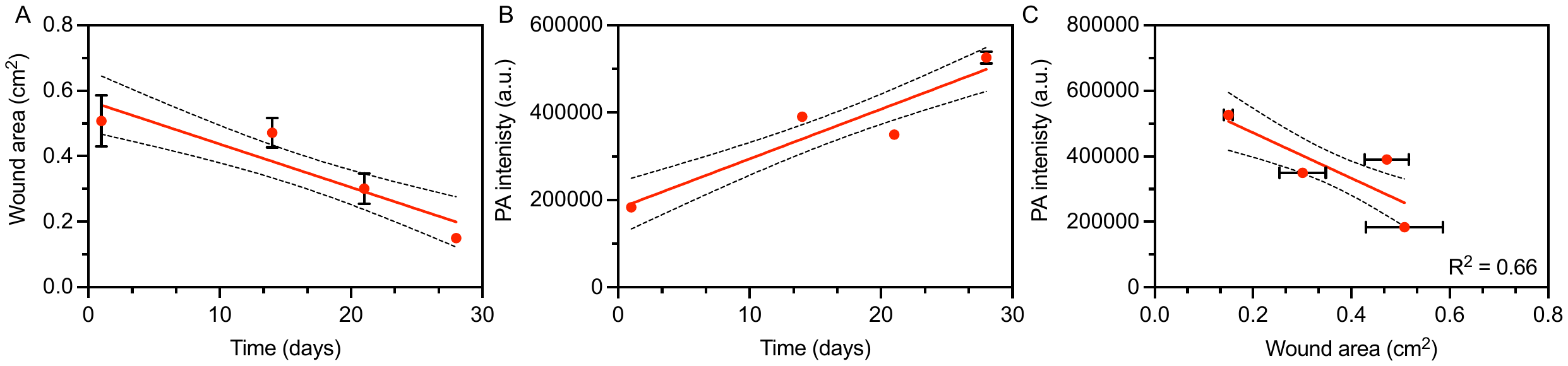

**Figure S24. A,** Wound area *vs.* time, **B**, PA intensity *vs.* time, and **C**, PA intensity *vs.* Wound area for PN17.

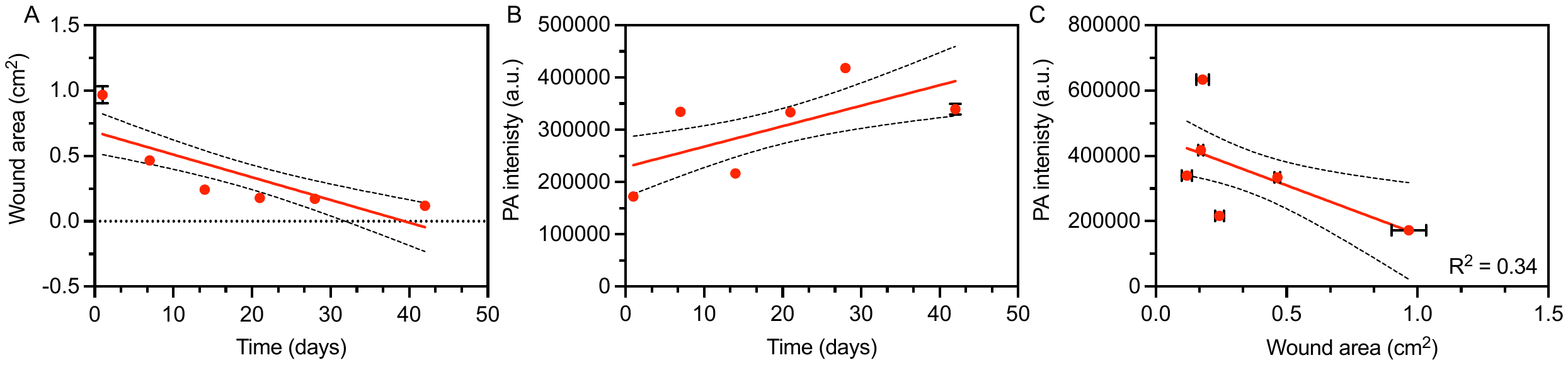

**Figure S25. A,** Wound area *vs.* time, **B**, PA intensity *vs.* time, and **C**, PA intensity *vs.* Wound area for PN18.

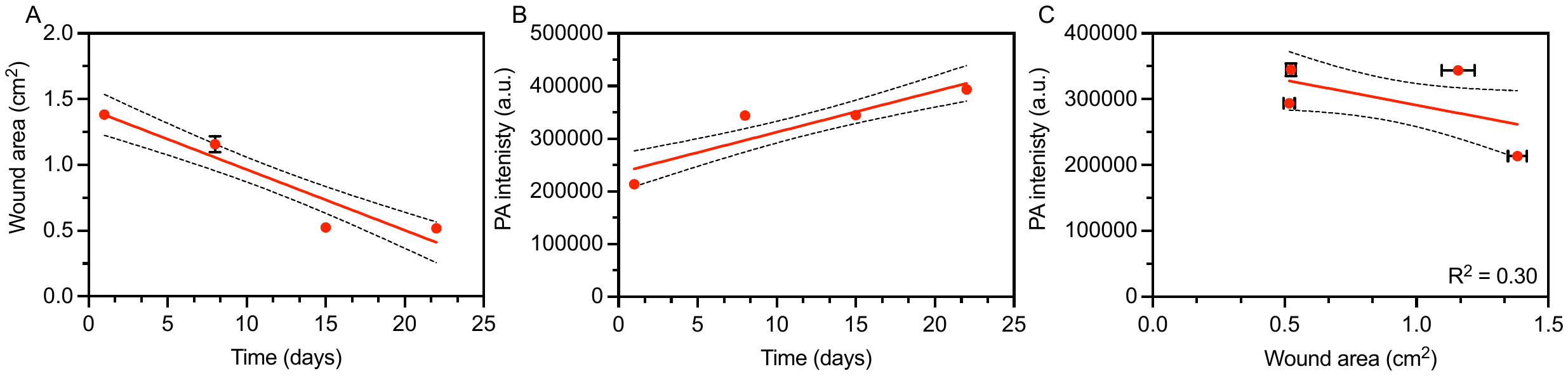

**Figure S26. A,** Wound area *vs.* time, **B**, PA intensity *vs.* time, and **C**, PA intensity *vs.* Wound area for PN19.

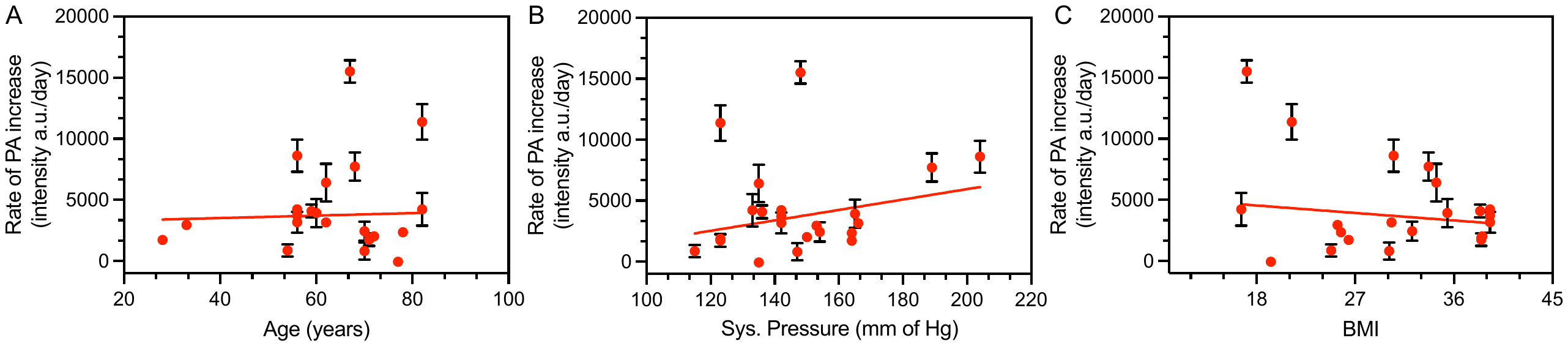

**Figure S26. A,** Effect of age (years), **B**, systolic blood pressure (mm of Hg), and **C**, body mass index (BMI) for the entire cohort on the rate of PA change shows no significant correlation.

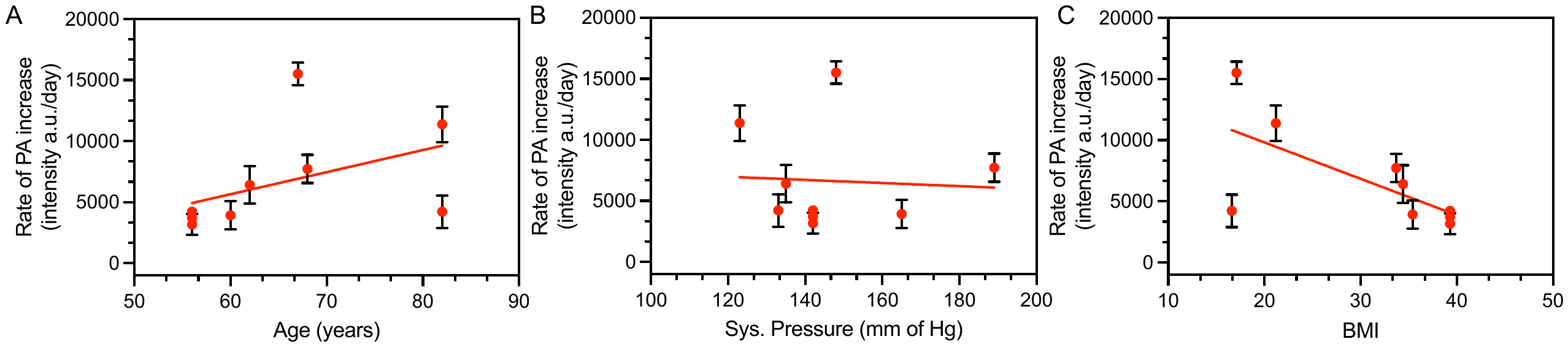

**Figure S27. A,** Effect of age (years), **B**, systolic blood pressure (mm of Hg), and **C**, body mass index (BMI) for the responders on the rate of PA change. Higher age and lower BMI are related to increased rate of change in PA intensity. Blood pressure had no significant effect on the rate of PA change.
